# The structure and organization of headache differential diagnoses: A Pilot Study of Subset Relationships between Differentials in ICHD3

**DOI:** 10.1101/2021.11.01.21265723

**Authors:** Pengfei Zhang

## Abstract

**Objective:** Differential diagnosis is fundamental to medicine. Using DiffNet, a differential diagnosis generator, as a model we studied the structure and organization of how collections of diagnose (i.e. sets of diagnoses) are related in the ICHD3. Specifically, we asked: Which sets of differential diagnoses are subsets of each other? What is the minimum number of sets of differential diagnoses that encompass all ICHD3 codes? Furthermore, we explored the clinical and theoretical implication of these answers.

**Methods:** DiffNet is a freely distributed differential diagnosis generator for headaches using graph theoretical properties of ICHD3. For each ICHD3 diagnosis, we generated a set of differential diagnoses using DiffNet. We then determined algorithmically the set/subset relationship between these sets. We also determined the smallest list of ICHD3 diagnosis whose differential diagnoses would encompass the totality of ICHD3 diagnoses.

**Results:** All ICHD3 diagnoses can be represented by a minimum of 92 differential diagnosis sets. Differential diagnosis sets for 10 of the 14 first digit subcategories of ICHD3 are represented by more than one differential diagnosis sets. Fifty-one of the 93 differential diagnosis sets contain multiple subset relationships; the remaining 42 do not enter into any set/subset relationship with other differential diagnosis sets. Finally, we included a hierarchical presentation of differential diagnosis sets in ICHD3 according to DiffNet.

**Conclusion:** We propose a way of interpreting headache differential diagnoses as partial ordered sets (i.e. poset). For clinicians, fluency with the 93 diagnoses and their differential put forth here implies a complete description of ICHD3. On a theoretical level, interpreting ICHD3 differential diagnosis as poset, allows researchers to translate differential diagnoses sets topologically, algebraically, and categorically.

## Introduction

Differential diagnosis is fundamental to clinical medicine.^1, 2^ Since candidate diagnoses in a differential form a collection of objects, this collection satisfies the mathematical definition of a set. To our knowledge, a project studying differential diagnosis hierarchically using set/subset relationship between sets of differentials has not been done. We sought to explore precisely the structure and organization of this hierarchy in headaches.

Although the International Classification of Headache Disorders (ICHD3) provides structure for headaches classifications, it does not inherently classify differential diagnoses. Nevertheless, the ICHD3 do provide differential diagnosis considerations by cross-references of diagnosis codes. For example, “headaches secondary to TIA” is not hierarchically associated wtih migraine with aura but the latter is a differential diagnosis of the former; this can be seen by the observation that migraine with aura (ICHD code 1.2) is listed as a reference in the comment section of headache caused by TIA (6.1.2). We took advantage of this observation in our previous work to construct DiffNet, a freely distributed differential diagnosis generator published under Rutgers University.^3, 4^ DiffNet is an image of ICHD3 in a graph theoretic form where crosses referenced of ICHD3 are interpreted as “differentials”.

Using DiffNet as a model, we asked: What is the set/subset relationship between differential diagnosis? What is the minimum sets of differential diagnosis sets needed to cover all of ICHD3 diagnosis codes? In answering these questions, we sought to provide a hierarchical classification and ordering of sets of differential diagnosis as a reference for clinicians. We believe such a classification would be invaluable when expanding/narrowing one’s differential diagnosis in headache. This arrangement of set/subset relationship also contains theoretical implications; for the technical readers, we explored these implications in Addendum 1.

## Methodology

Our study was conducted in two phases: 1) the differential diagnosis generation phase and 2) the set/subset generation phase. The differential diagnosis generation phase consisted of the building of DiffNet; we will review it here.^4^ The set/subset generation phase involved determining the set/subset relationships between differential diagnosis groupings.

### The Building of DiffNet, the differential diagnosis generator

DiffNet reinterprets ICHD3 as a graph in the following fashion: each ICHD3 diagnosis is considered a “node”; if one ICHD3 diagnosis references another ICHD3 diagnosis anywhere in the criterion or comment section, then an “edge” exists between the two.

Furthermore, an edge exists between two ICHD3 diagnoses if they are connected through the ICHD3 hierarchy.

For example 6.1 and 6.1.1 are connected since the latter is a sub-section diagnosis of the former. However, 6.1.1.1 and 6.1.1.2 are not necessarily connected as an edge as they are on the same level. (On the other hand, 6.1 and 6.1.1.1 are connected despite that they are once removed.) This ICHD3 graph is undirected and contains 387 nodes (due to diseases with duplicate diagnostic codes, for example “complications of migraine”, 1.4 and A1.4). An editorial choice was made by DiffNet to consider duplicated diagnosis the same as long as they are called by the same name; for example 1.4 and A1.4 are considered as the same entity.

DiffNet then operates in the following fashion to generate differential diagnosis: when given an ICHD3 diagnosis code as an input node, DiffNet look for all “first-degree neighbors” of that diagnostic code. First-degree neighbor is graph theoretic parlance for all nodes connected to a given node via an edge. These “first degree neighbors” are output to the user as differential diagnosis. (DiffNet also provides “second degree neighbors” as well in order to describe less likely differential diagnoses. We ignore these in this project.)

Consider “cervicogenic headache” as an example: The “first degree neighbors” of this node/ICHD3 diagnosis are the following:

headache attributed to cervical myofascial pain
headache attributed to upper cervical radiculopathy
headache attributed to disorder of the neck
migraine
chronic tension-type headache associated with pericranial tenderness
infrequent episodic tension-type headache associated with pericranial tenderness
frequent episodic tension-type headache associated with pericranial tenderness
tension-type headache (tth)
headache or facial pain attributed to disorder of the cranium neck eyes ears nose sinuses teeth mouth or other facial or cervical structure
diving headache

In other words, the above are ICHD3 diagnoses that are either mentioned in the criterion/comment section – such as “Headache attributed to cervical myofascial pain” (A11.2.5) - or is connected via ICHD3 hierarchy – such as, “Headache attributed to disorder of the neck”(11.2). Notice that edges that are established elsewhere in ICHD3 are also included in the differential. For example, “diving headache” is included since it was mentioned not in the cervicogenic headache section but rather in 10.1.3 as a potential differential of diving headache: “… 11.2.1 cervicogenic headache can occur during a dive.” (page 140 of ICHD3).^5^

The building of DiffNet was accomplished through Python with NetworkX library; C# was used as an interface. The algorithm is freely distributed through user registration by Rutgers University at http://license.rutgers.edu/technologies/2019-070_diffnet-headache-differential-diagnosis-software. (To use algorithm, simply enter input diagnosis as lower case.) The source code is proprietary to Rutgers University but maybe opened to limited disclosure with inquiry to the author. The parsing of ICHD3 for DiffNet was accomplished through the Haskell programming language.

### Determining Set/Subset Relationships

Once DiffNet was generated, each of the ICHD3 diagnoses code was algorithmically entered into DiffNet to generate a list of differential. We call the set of differential diagnosis generated by a specific ICHD3 diagnostic node the differential diagnosis set *induced* by that specific diagnostic code. For example, in our example above, the set of differential induced by “cervicogenic headache” is the first-degree neighbors generated by DiffNet.

Once sets of differential diagnoses induced by each of the ICHD3 diagnostic codes were obtained, we then algorithmically obtained all possible *proper subset* relationships between differential diagnosis sets.

Recall that subset and proper subset is defined mathematically as the following:

Let A and B be sets, B is a subset of A if and only if every member of B is a member of A. We call A the super-set of B. Furthermore, B is a proper subset of A if it is strictly contained in A.^6–8^

Consider the following example on three sets. The differential diagnosis set induced by “migraine with aura” is:

familial hemiplegic migraine type 3 (fhm3)
typical aura with headache
familial hemiplegic migraine type 1 (fhm1)
pure menstrual migraine with aura
probable migraine
chronic migraine (alternative criteria)
headache attributed to moyamoya angiopathy (mma)
episodic syndromes that may be associated with migraine
sporadic hemiplegic migraine (shm)
benign paroxysmal torticollis
headache attributed to mitochondrial encephalopathy lactic acidosis and stroke-like episodes (melas)
angiography headache
headache attributed to an intracranial endarterial procedure
familial hemiplegic migraine (fhm)
triptan-overuse headache
menstrually related migraine with aura
familial hemiplegic migraine type 2 (fhm2)
headache attributed to other chronic intracranial vasculopathy
migraine aura status
headache attributed to transient ischaemic attack (tia)
status migrainosus
vestibular migraine
typical aura without headache
migraine with typical aura
migraine aura-triggered seizure
familial hemiplegic migraine other loci
retinal migraine
visual snow
migraine with brainstem aura infantile colic
chronic migraine hemiplegic migraine
persistent aura without infarction
headache attributed to cerebral autosomal dominant arteriopathy with subcortical infarcts and leukoencephalopathy (cadasil)
probable migraine with aura
headache attributed to arteriovenous malformation (avm)
migrainous infarction
non-menstrual migraine with aura
migraine without aura
headache attributed to cerebral venous thrombosis (cvt)
migraine
migraine with aura

The differential diagnosis set induced by “familial hemiplegic migraine (fhm)” is:

familial hemiplegic migraine type 3 (fhm3)
familial hemiplegic migraine type 1 (fhm1)
hemiplegic migraine
familial hemiplegic migraine (fhm)
sporadic hemiplegic migraine (shm)
familial hemiplegic migraine other loci
familial hemiplegic migraine type 2 (fhm2)
migraine
migraine with aura

Furthermore differential diagnoses induced by “familial hemiplegic migraine type 3 (fhm3)” is:

familial hemiplegic migraine type 3 (fhm3)
migraine
familial hemiplegic migraine (fhm)
hemiplegic migraine
migraine with aura

We can write that the differential diagnosis set induced by “familial hemiplegic migraine type3” (we call this *differential set* for short) is a proper subset of differential set induced by “familial hemiplegic migraine (fhm)”, which in turn in a proper subset of differential set induced by “migraine with aura”. Symbolically, we can write the following: differential diagnosis set induced by “familial hemiplegic migraine type3” < differential set induced by “familial hemiplegic migraine (fhm)” < differential set induced by “migraine with aura”.

All such set/subset relationships between differential diagnosis sets in the ICHD3 were determined and recorded. (These relationships form a poset in order theory parlance. We investigated the theoretical implications of this in the technical addendum.)

We then algorithmically determined the minimum set of ICHD3 whose differential diagnoses encompass the totality of ICHD3. We call these sets the *lower-sets*. We further analyzed the resultant set/subset relationships by comparing them against the ICHD3 categories.

The set/subset phase of the project was accomplished through the Haskell programming language. Figure 1 was generated using Gephi. Codes are available upon request from the author with permission from Rutgers University.

**Fig 1.**
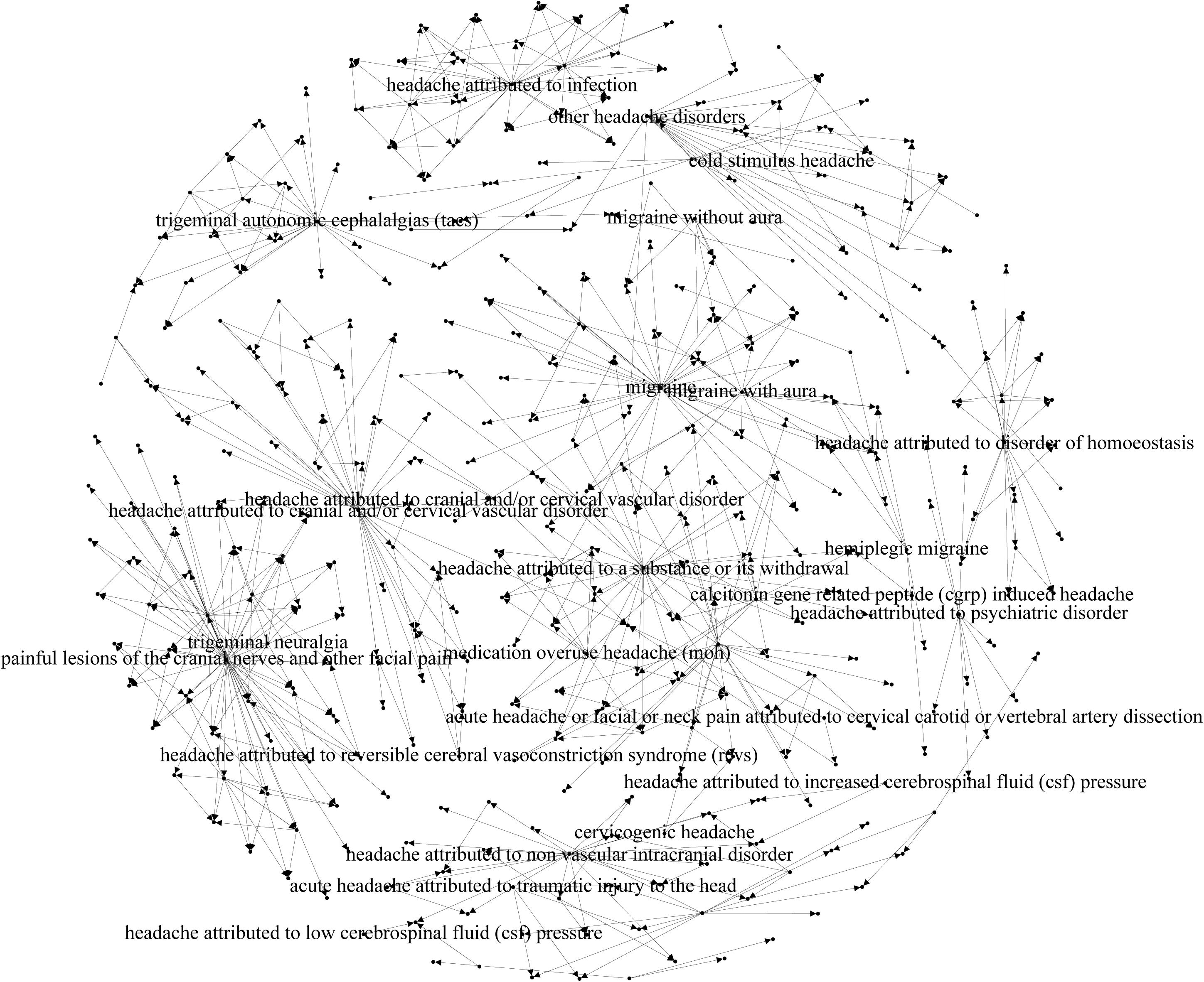

## Results

A total of 1368 edges and 387 nodes were generated by DiffNet. There are 584 proper subset relationships. The differentials encompass all of the ICHD3 diagnoses. Of these, all 14 first digit level of the ICHD3 diagnostic criteria are present.

Table 1 displays the 91 lower-sets, of these 51 contains subsets (Table 1a). There are 42 differential sets (we call them *singletons*) that are neither subset nor superset of another set. We display these in Table 1b. The complete lists of all set/subset relationships are displayed in Table 2, hierarchically, similar to ICHD3. We will reference each diagnosis by their section and subsection numbers. For example, “visual snow” is “31v”.

**Table 1a.**
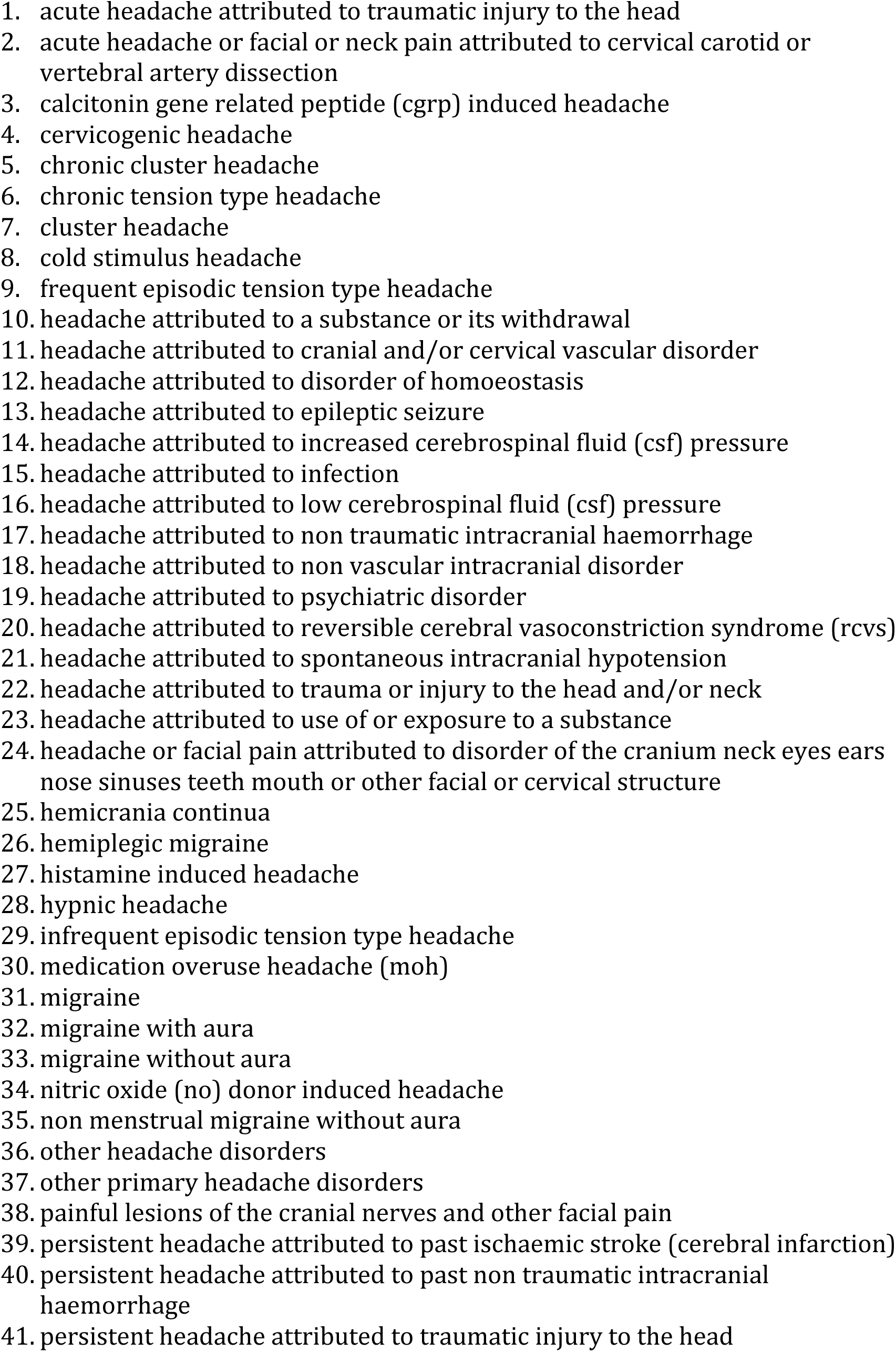
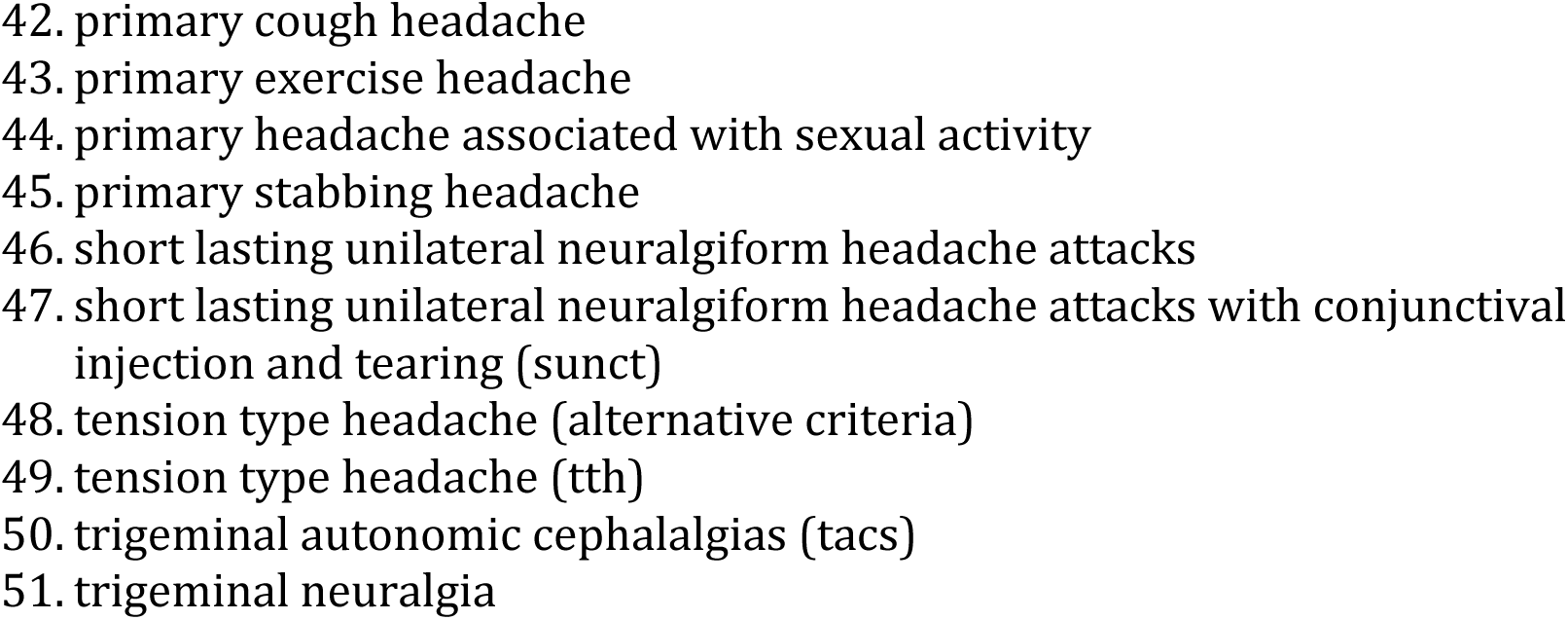
A list of Lower Sets

**Table 1b:**
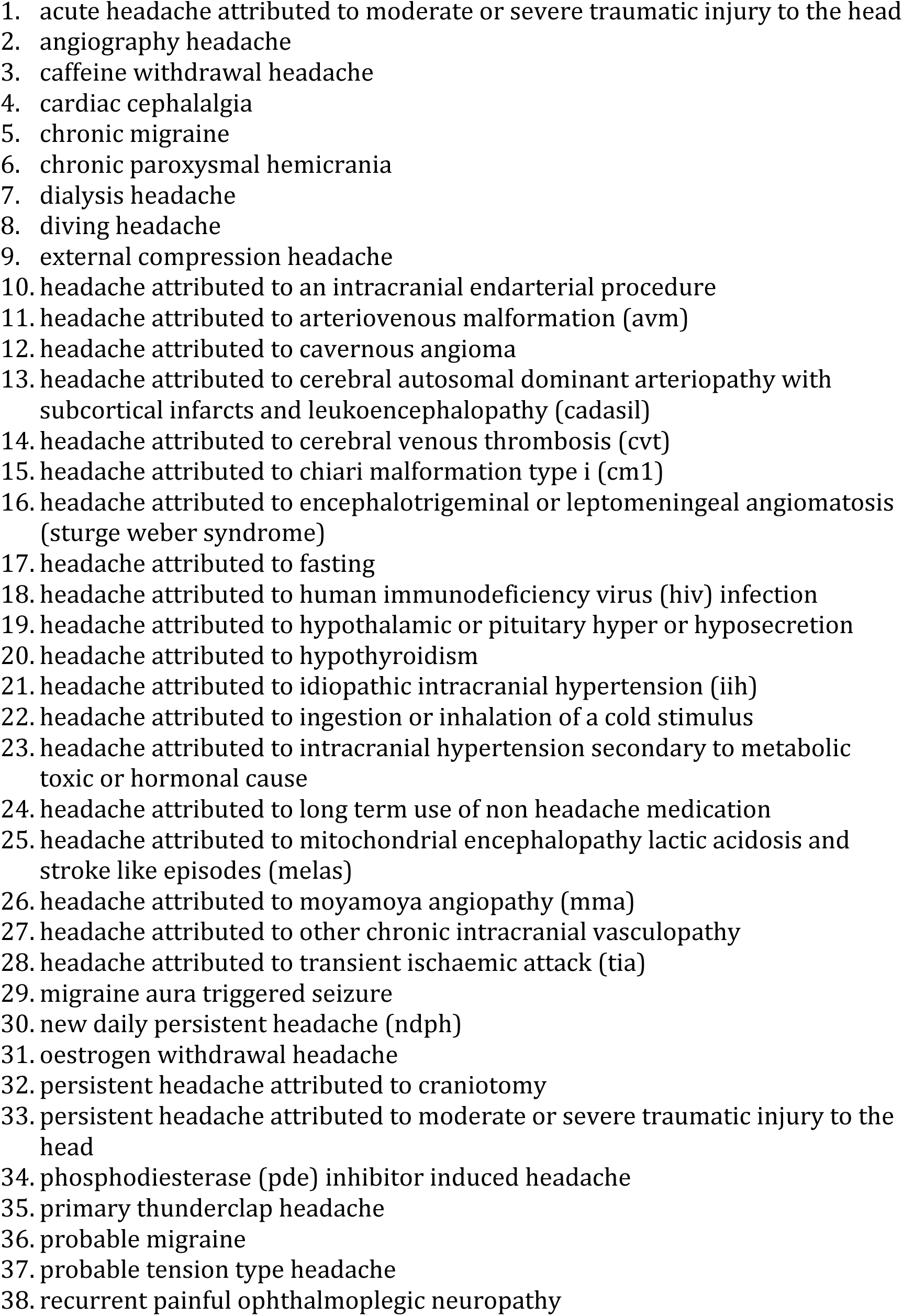
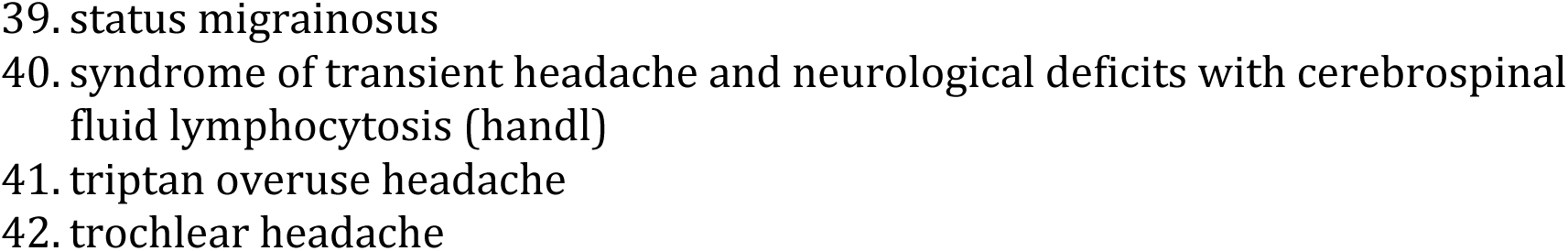
A list of Singletons

**Table 2.**
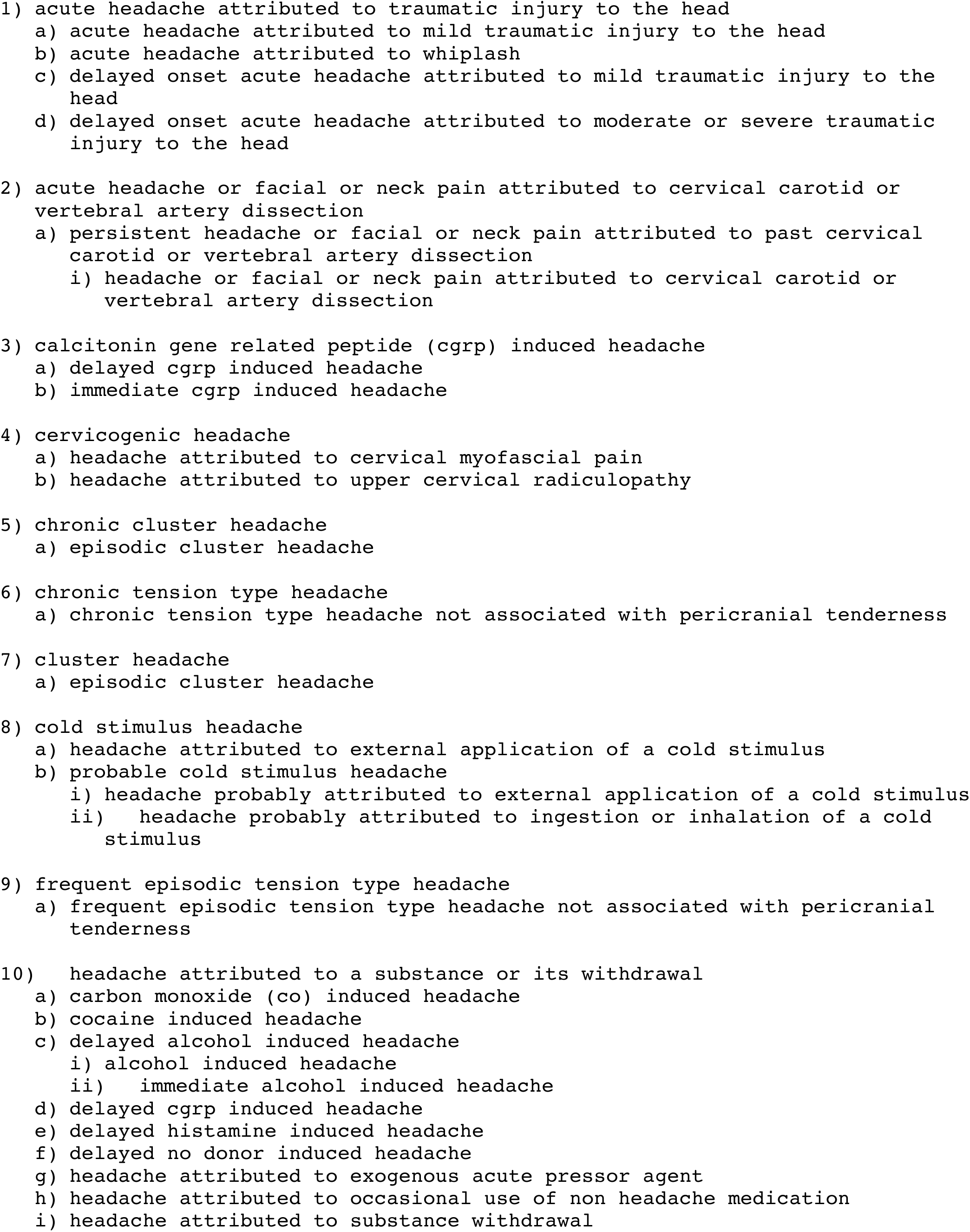
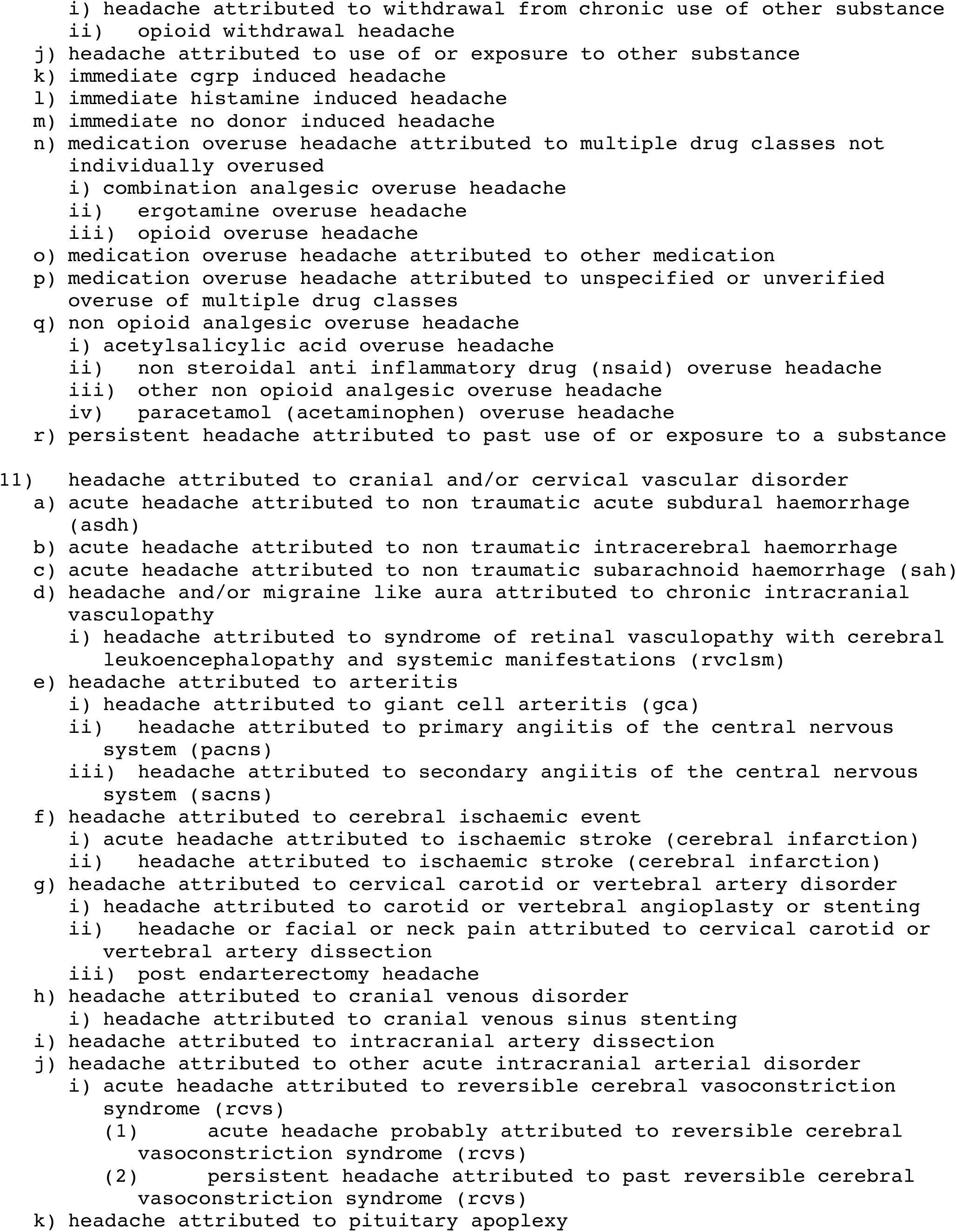
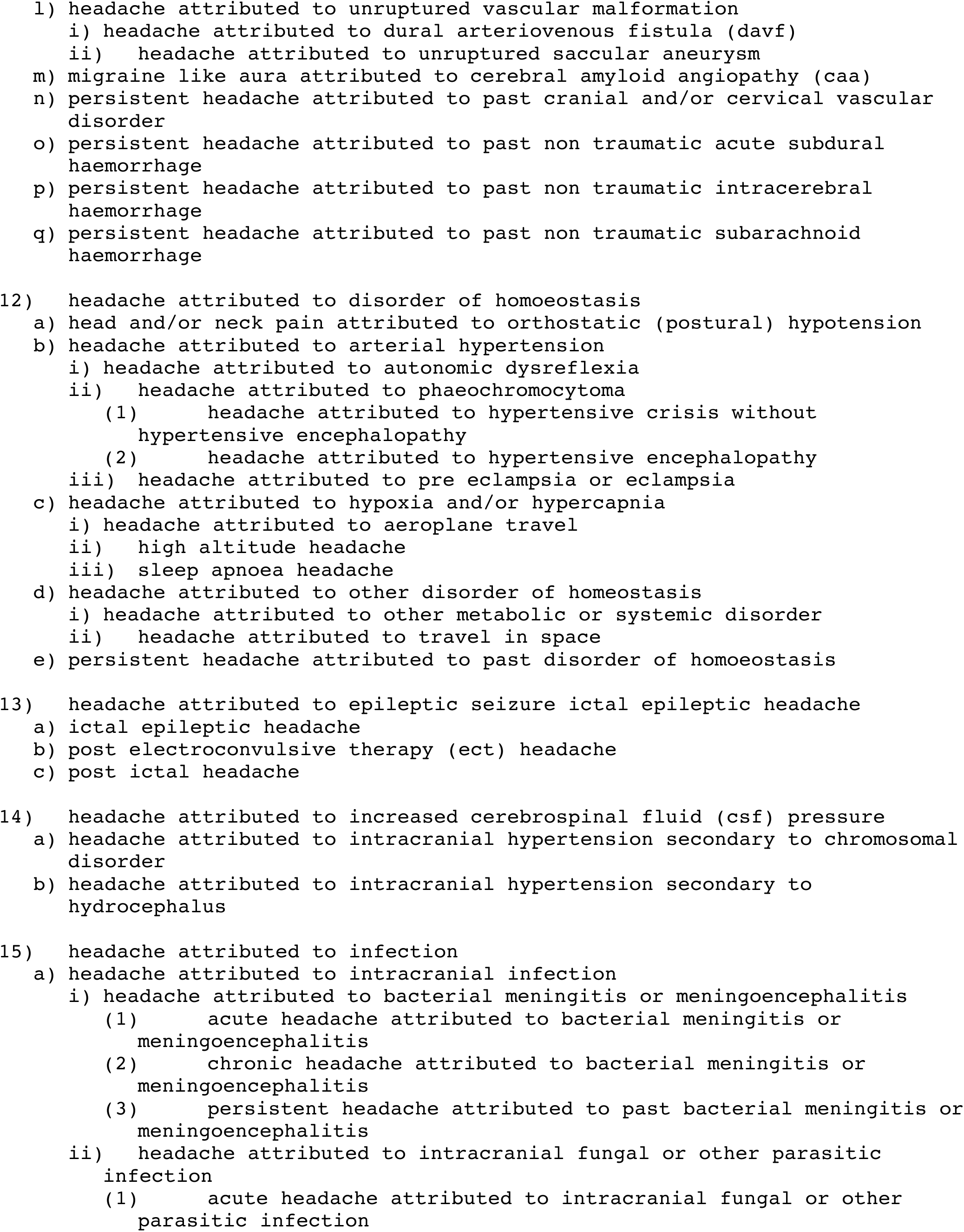
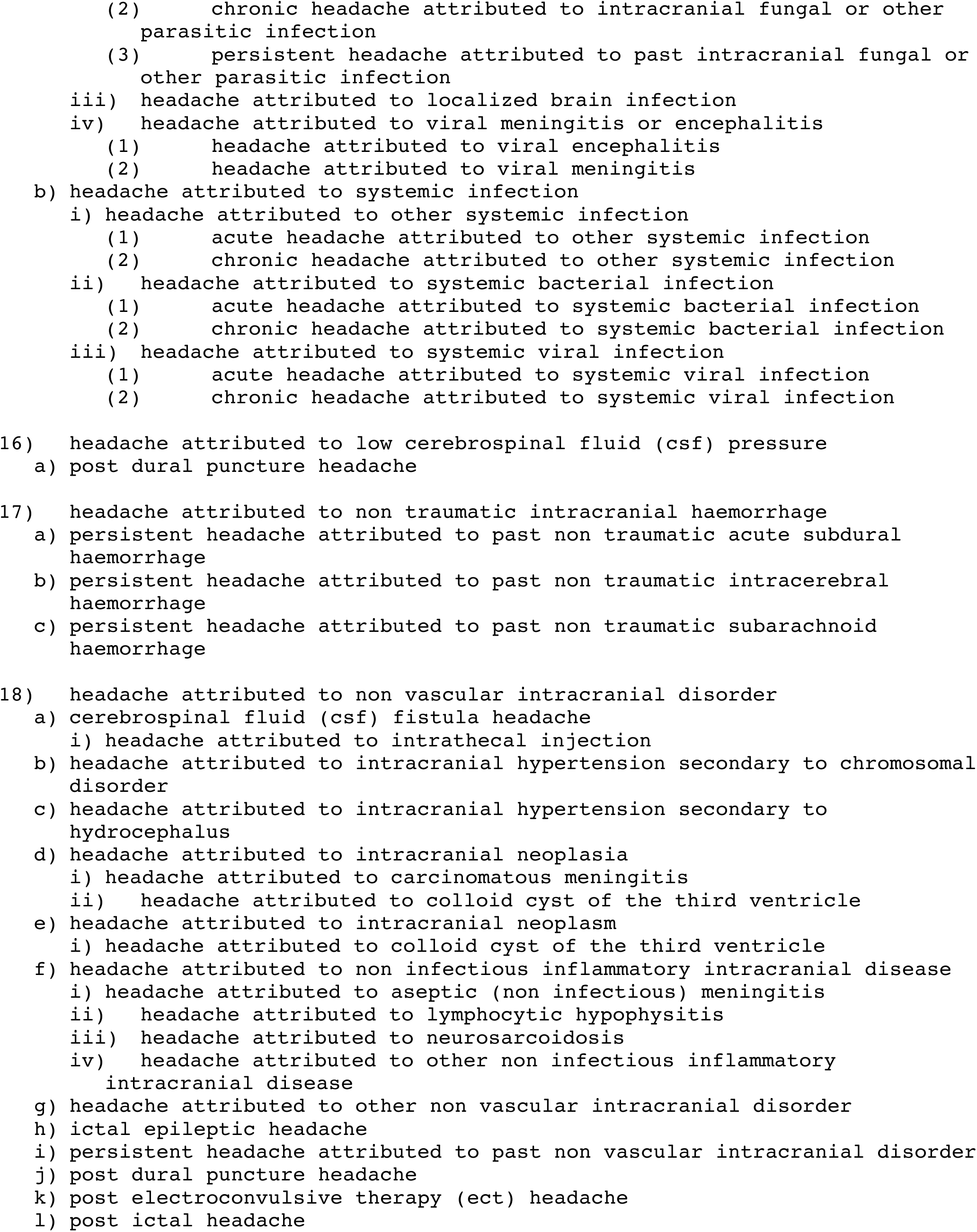
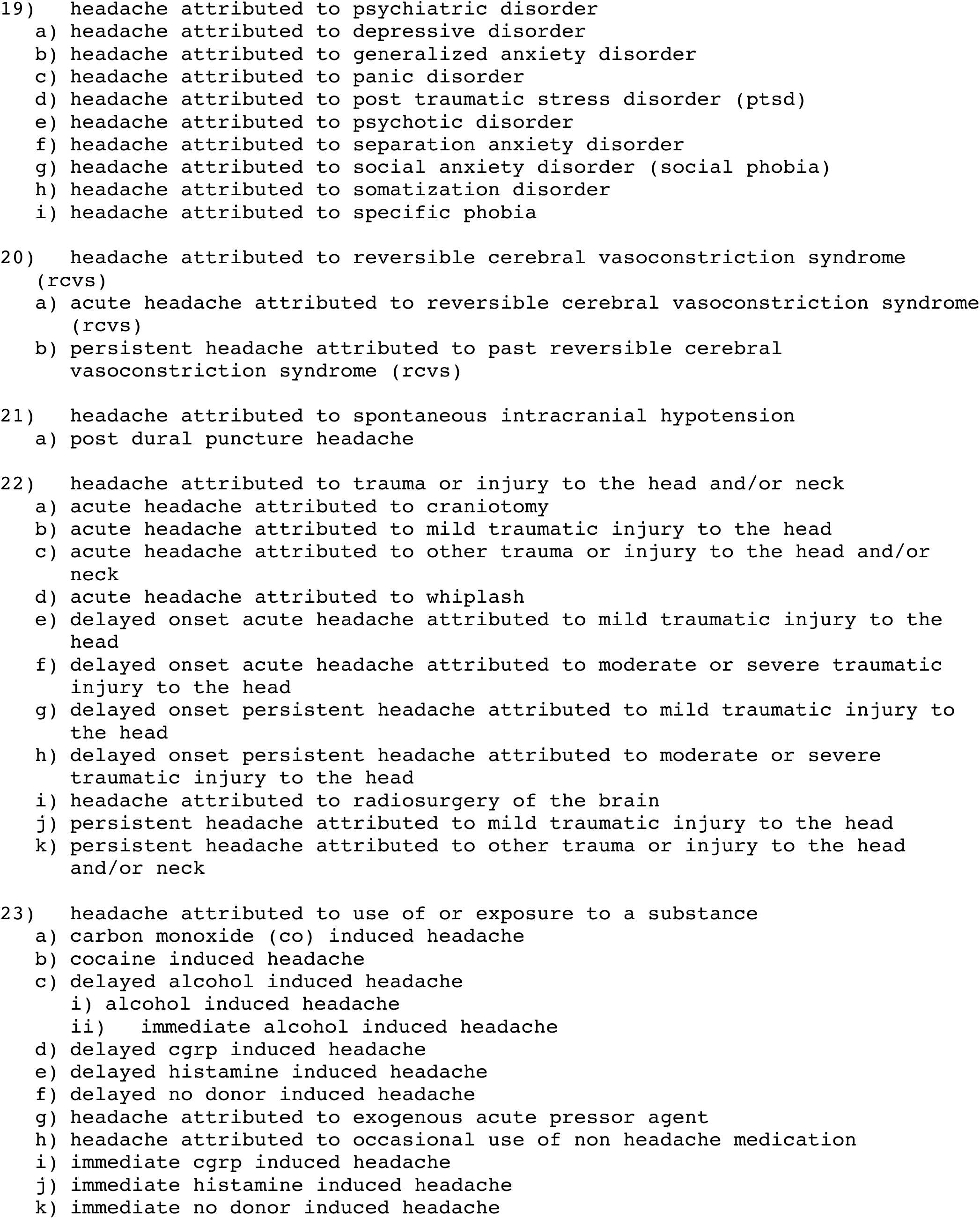
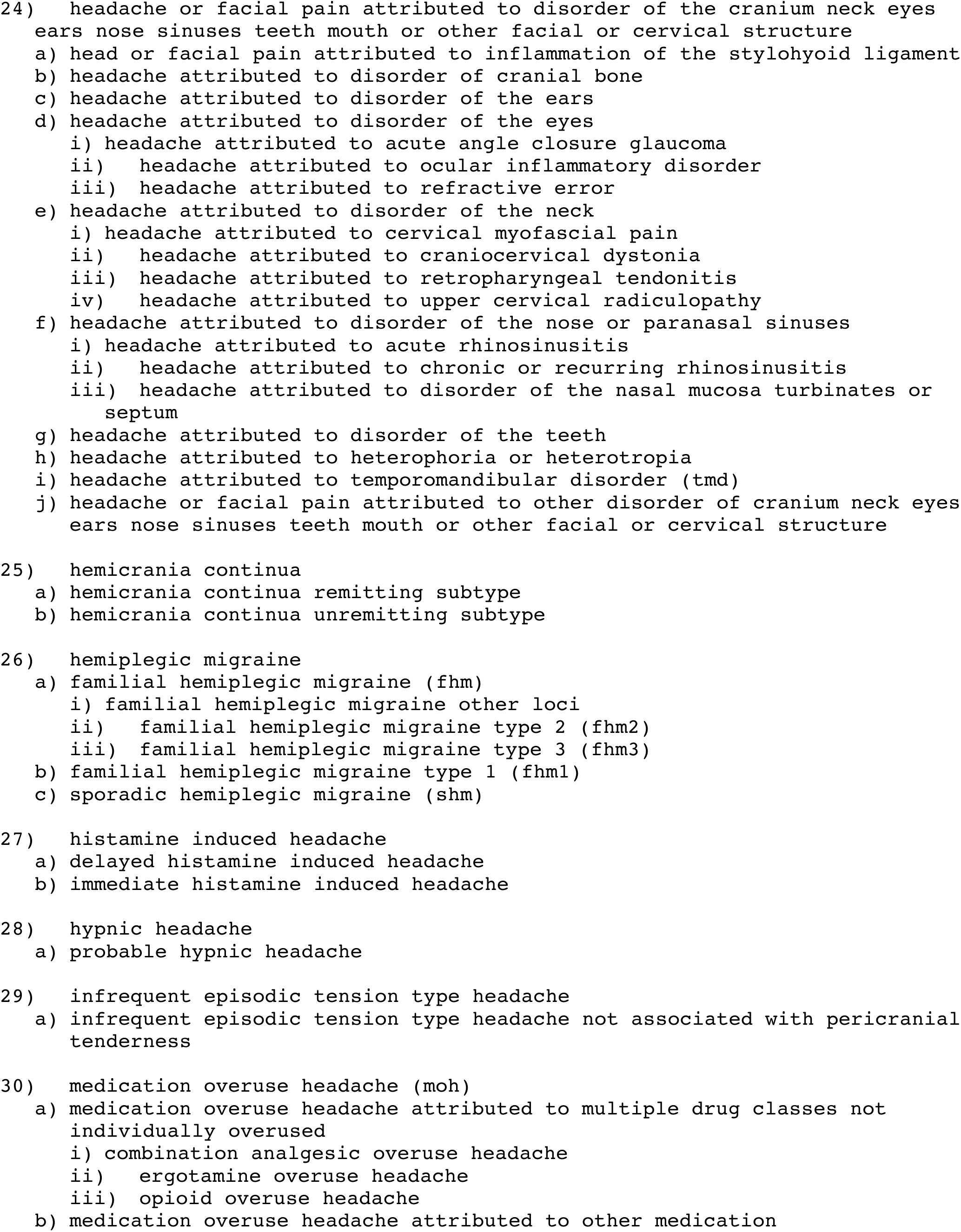
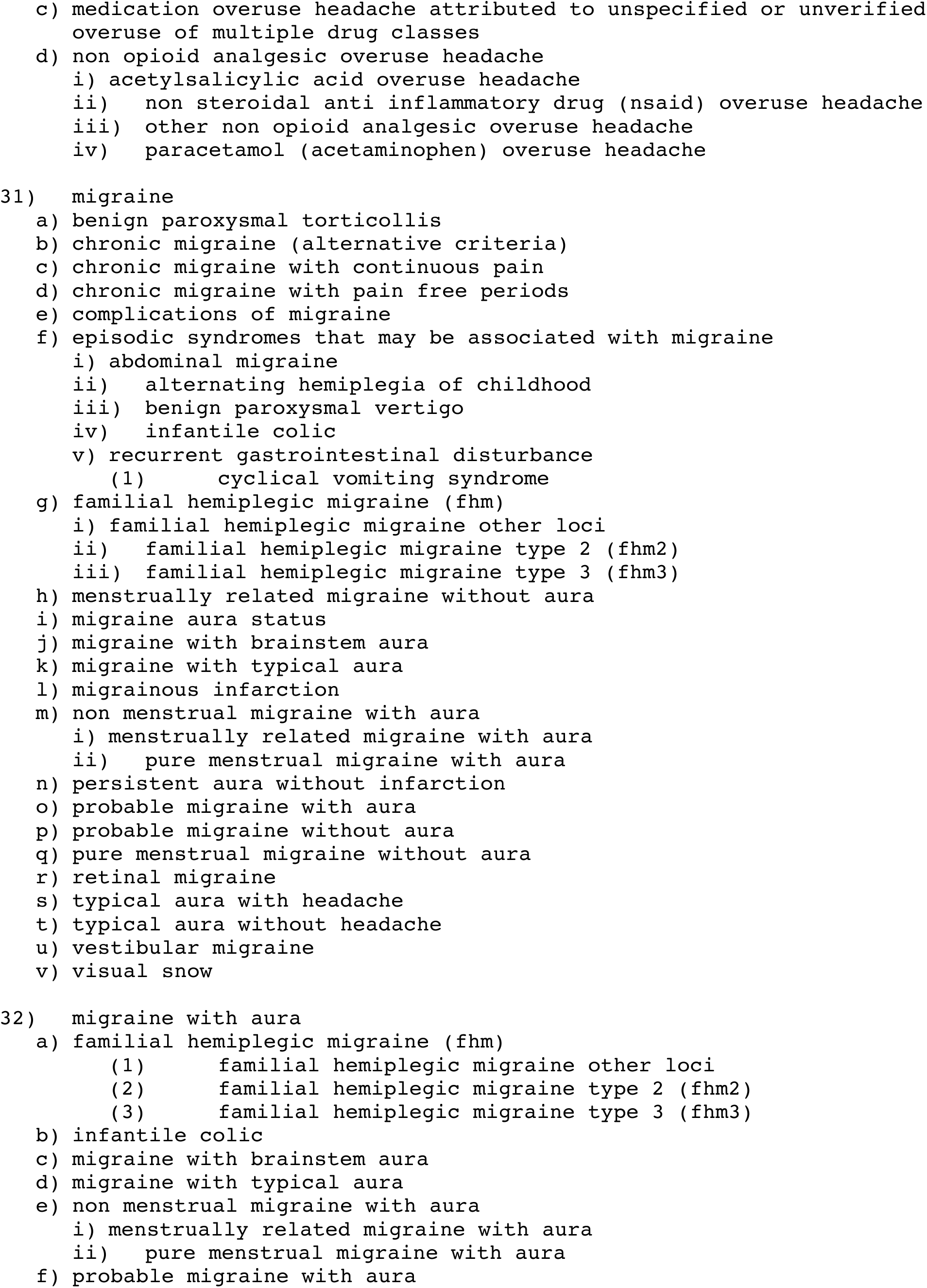
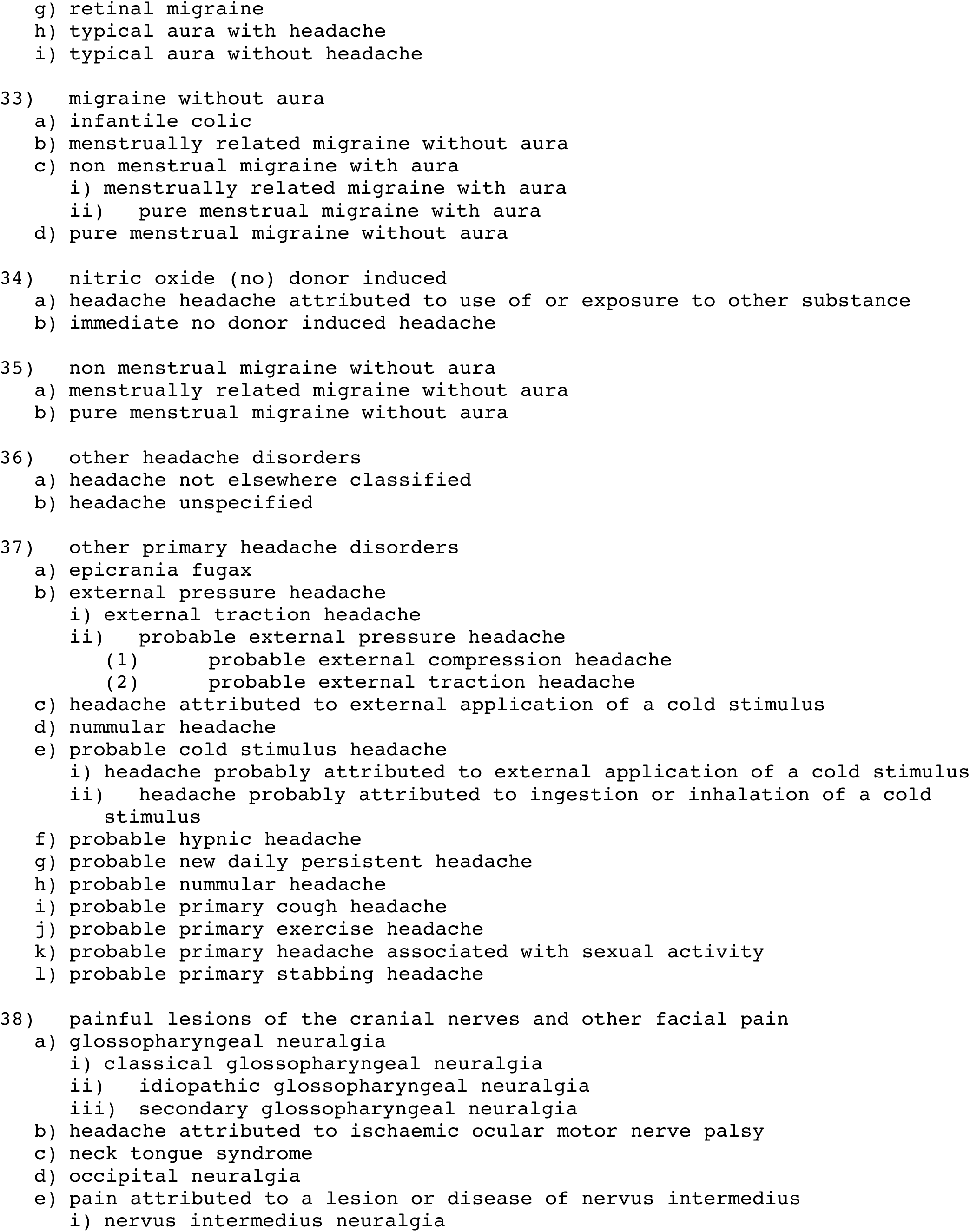
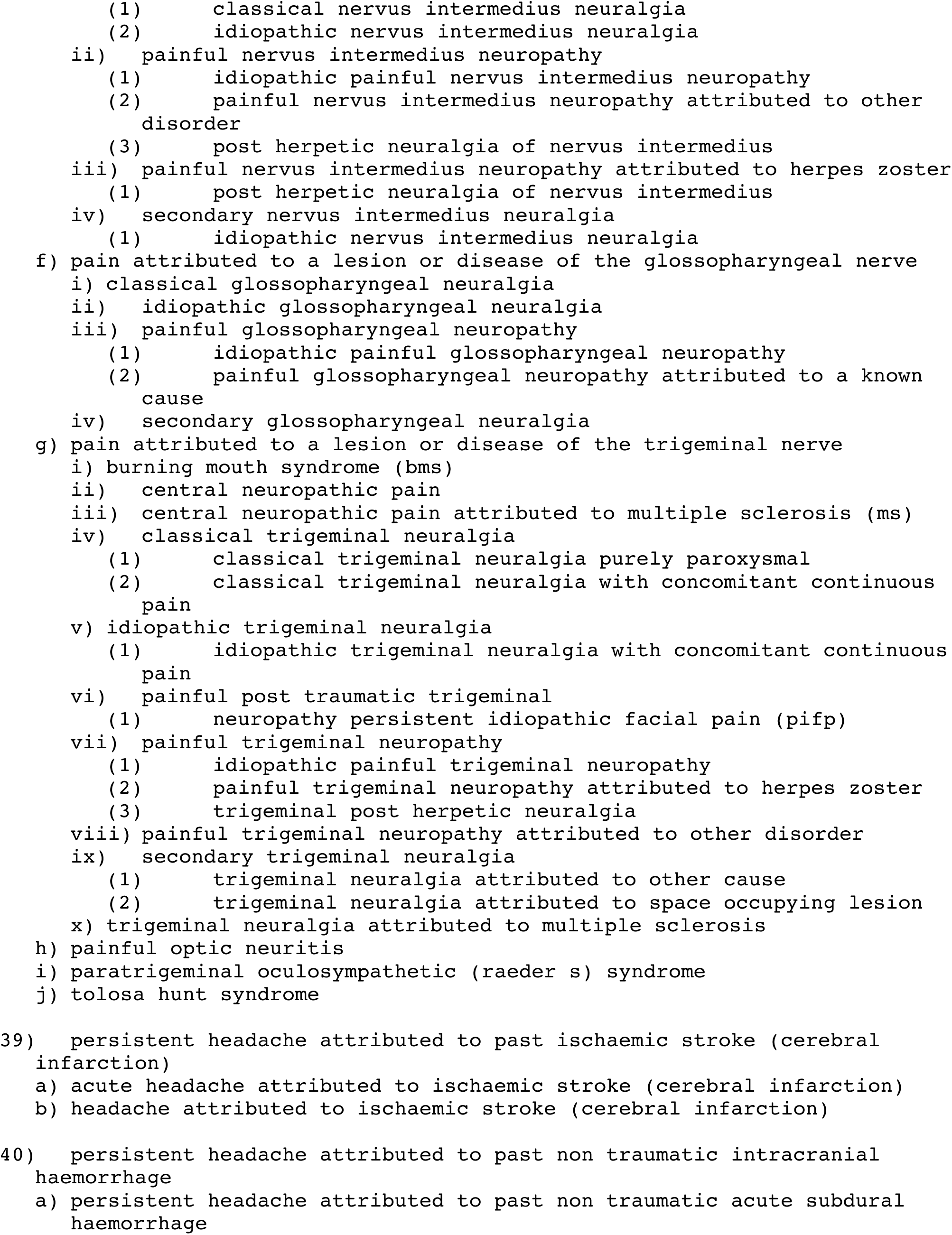
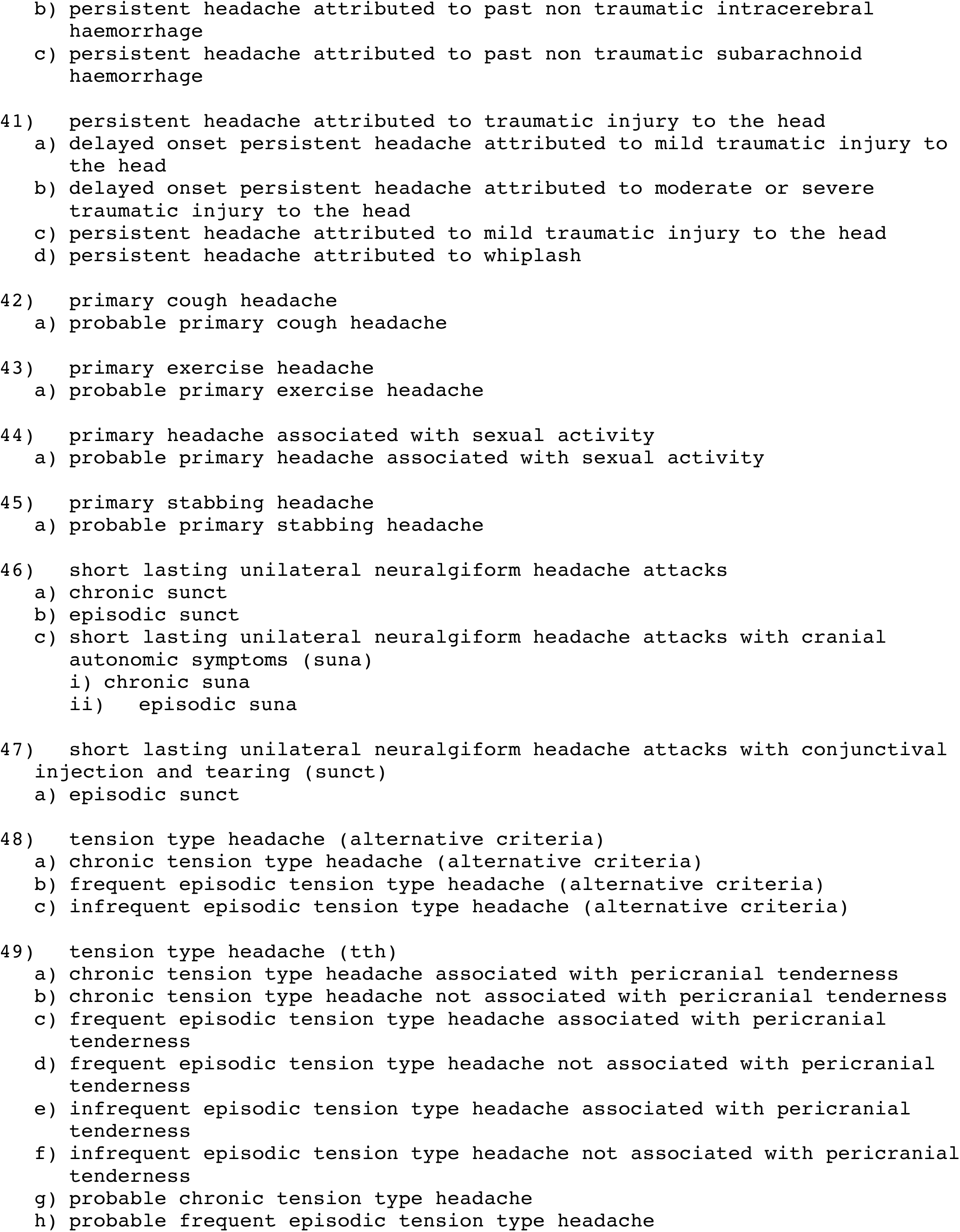
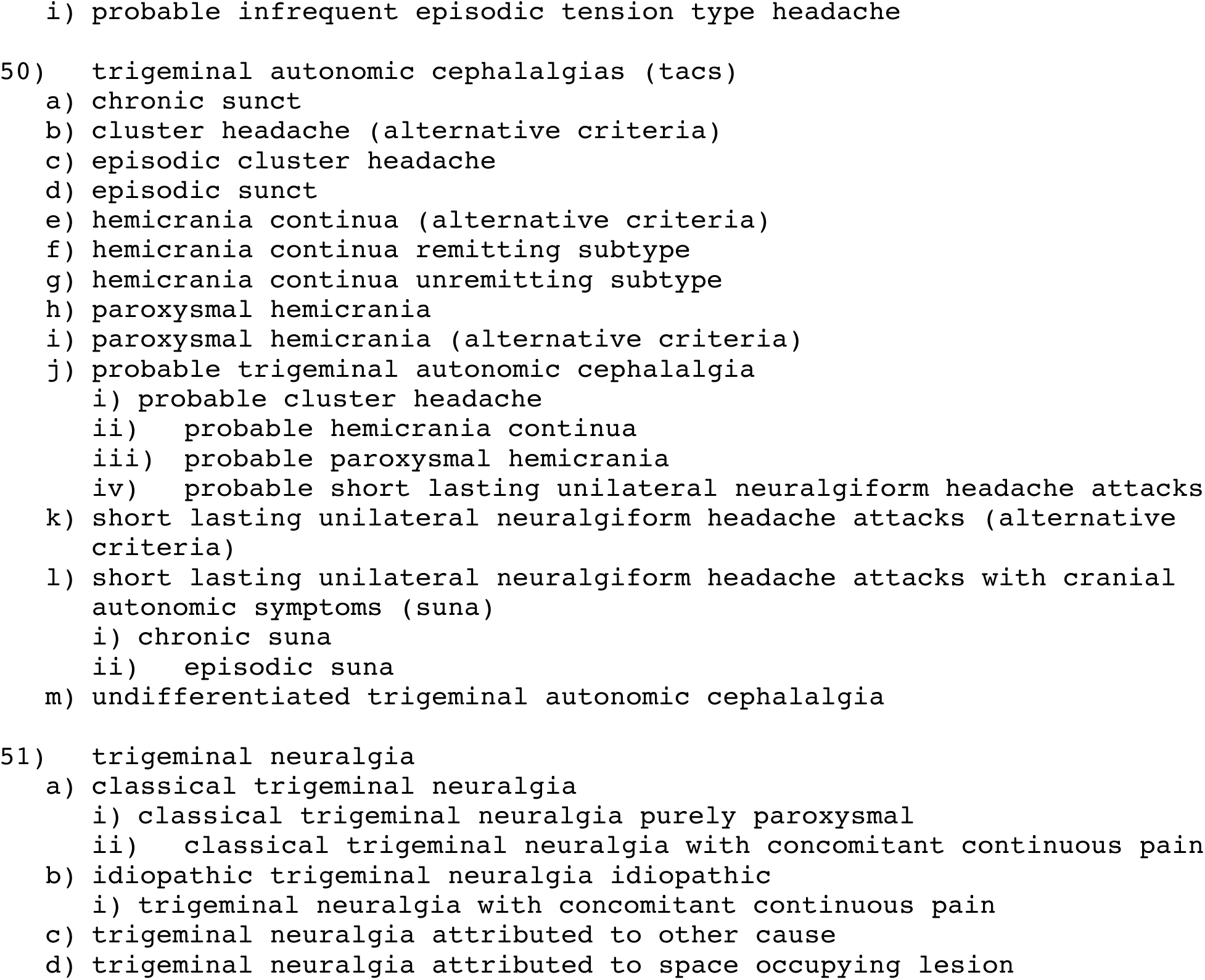

The 14 first digit levels of ICHD3 criteria are represented by the following differential groups:

1. Migraine is represented by sets 26, 31, 32, 33, 35.
2. Tension-type headaches is represented by sets 6, 9, 29, 48, 49, 50.
3. Trigeminal autonomic cephalalgias is represented by sets 5, 7, 25, 46, 47.
4. Other primary headache disorders is represented by sets 8, 28, 37, 42, 43, 44, 45.
5. Headache attributed to trauma or injury to the head and/or neck is represented by sets 1, 22, 41.
6. Headache attributed to cranial or cervical vascular disorder is represented by sets 2, 11, 17, 20, 39, 40.
7. Headache attributed to non-vascular intracranial disorder is represented by sets 13, 14, 16, 18, 21.
8. Headache attributed to a substance or its withdrawal is represented by sets 3, 10, 23, 27, 30, 34.
9. Headache attributed to infection is represented by sets 15.
10. Headache attributed to disorder of homeostasis is represented by sets 12.
11. Headache or facial pain attributed to disorders of the cranium, neck, eyes, ears, nose, sinuses, teeth, mouth or other facial or cervical structure is represented by sets 4, 24.
12. Headache attributed to psychiatric disorder is represented by sets 19.
13. Painful lesions of the cranial nerves and other facial pain is represented by sets 38, 51.
14. Other headache disorders is represented by sets 36.

We present the set/subset relationship graphically with arrows pointing from the super-set to its subset for visualization in Figure 1. We excluded the 42 singleton from this graph as these data points contain no arrow.

Twelve “inversions”/”violators” of ICHD3 hierarchical orders exist for differential sets. Consider two examples: A1.2 Migraine with Aura is hierarchically not connected to A1.6.4, infantile colic, but the differential set of the latter is a subset of the former. Diagnosis code 8.1.4.2 Delayed alcohol-induced headache is under 8.1.4 alcohol-induced headache, however the former is a superset for the latter. We present these in Table 3. Here, superset and subset relationships are represented by the format “superset, subset”.

**Table 3:**
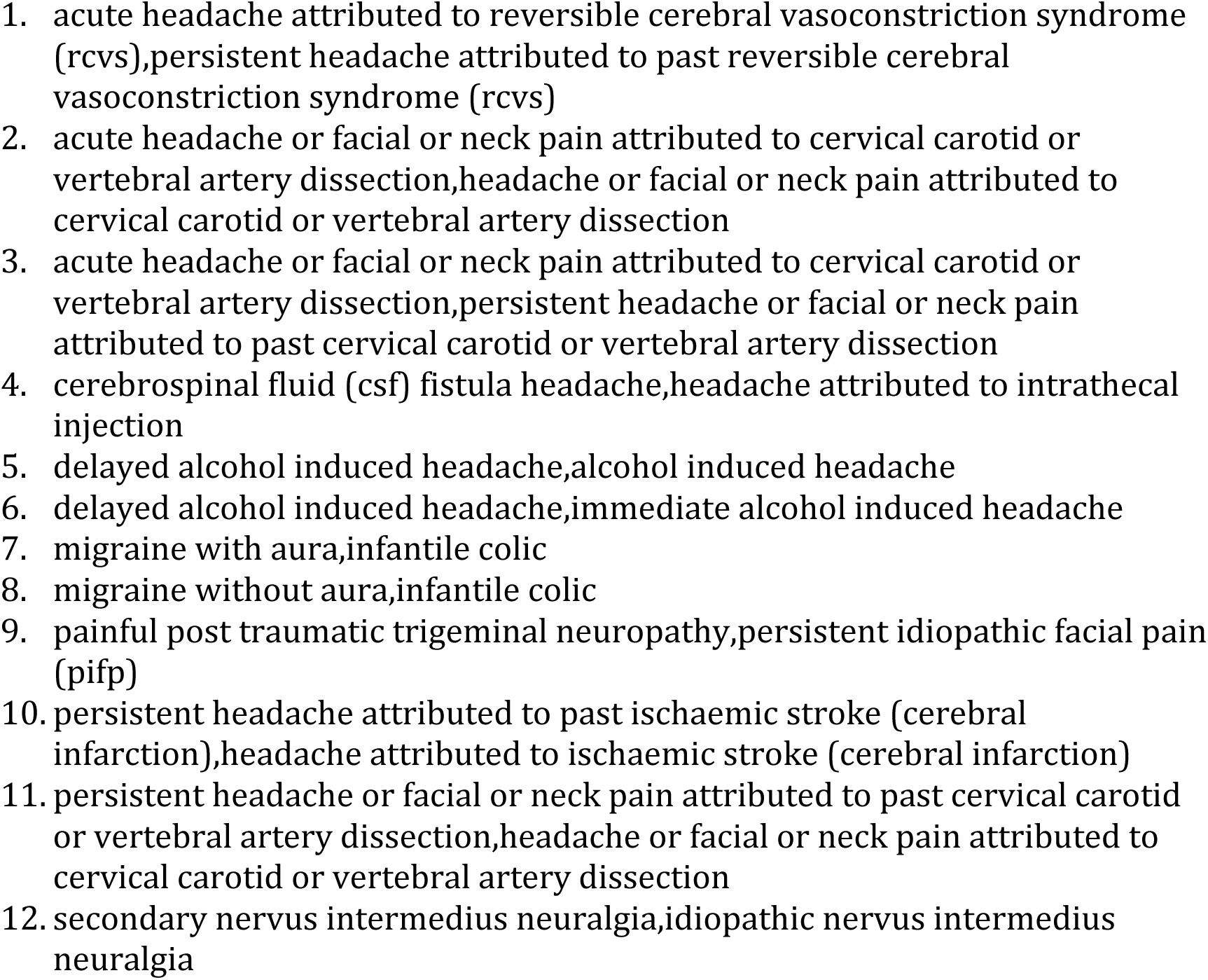
The inversions

Finally, the “lower bounds” (i.e. “meet”) of differential sets were calculated and presented in Table 4. A technical addendum (Addendum 1) is included to explore this and other theoretical implications of this project.

**Table 4.**
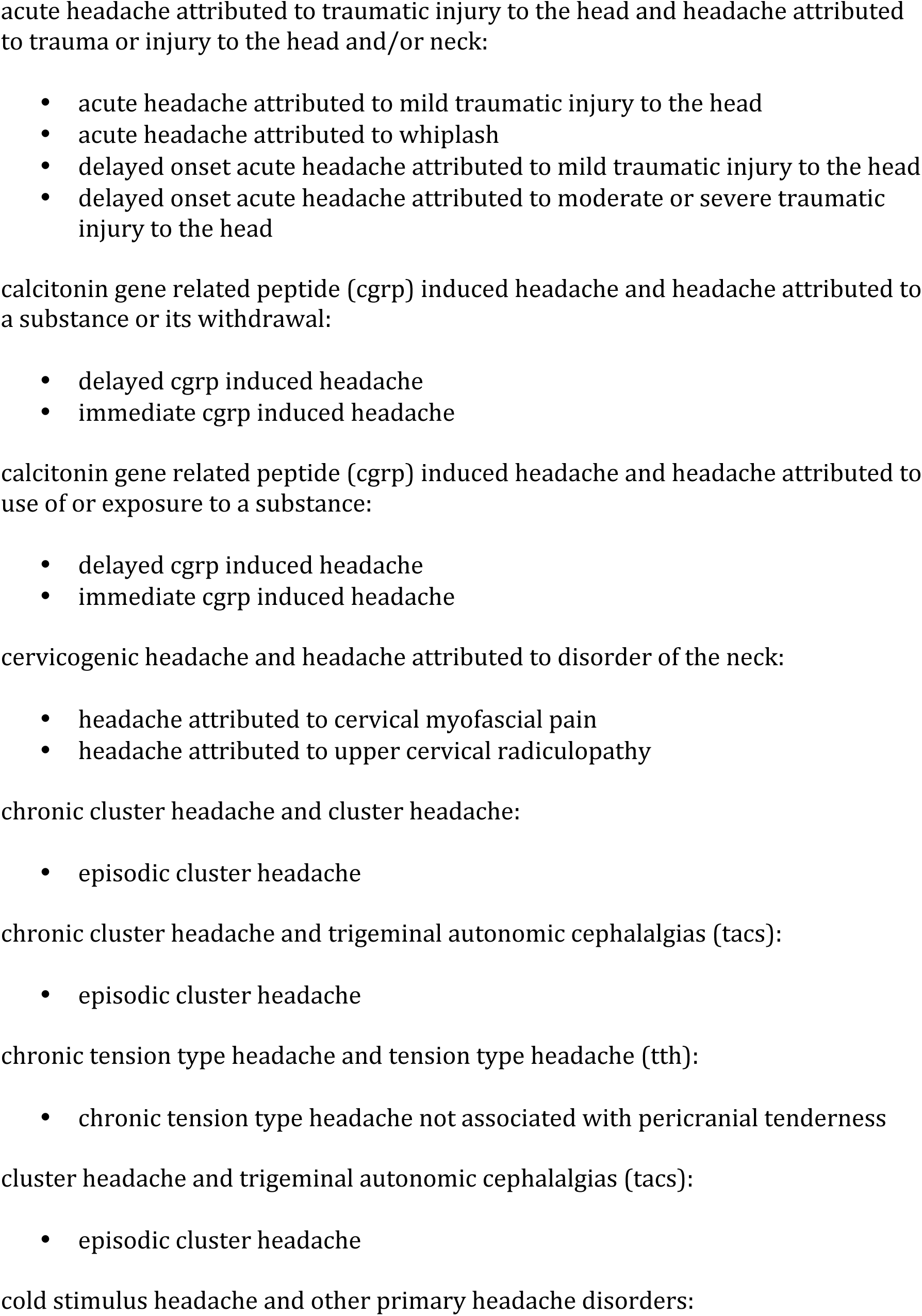
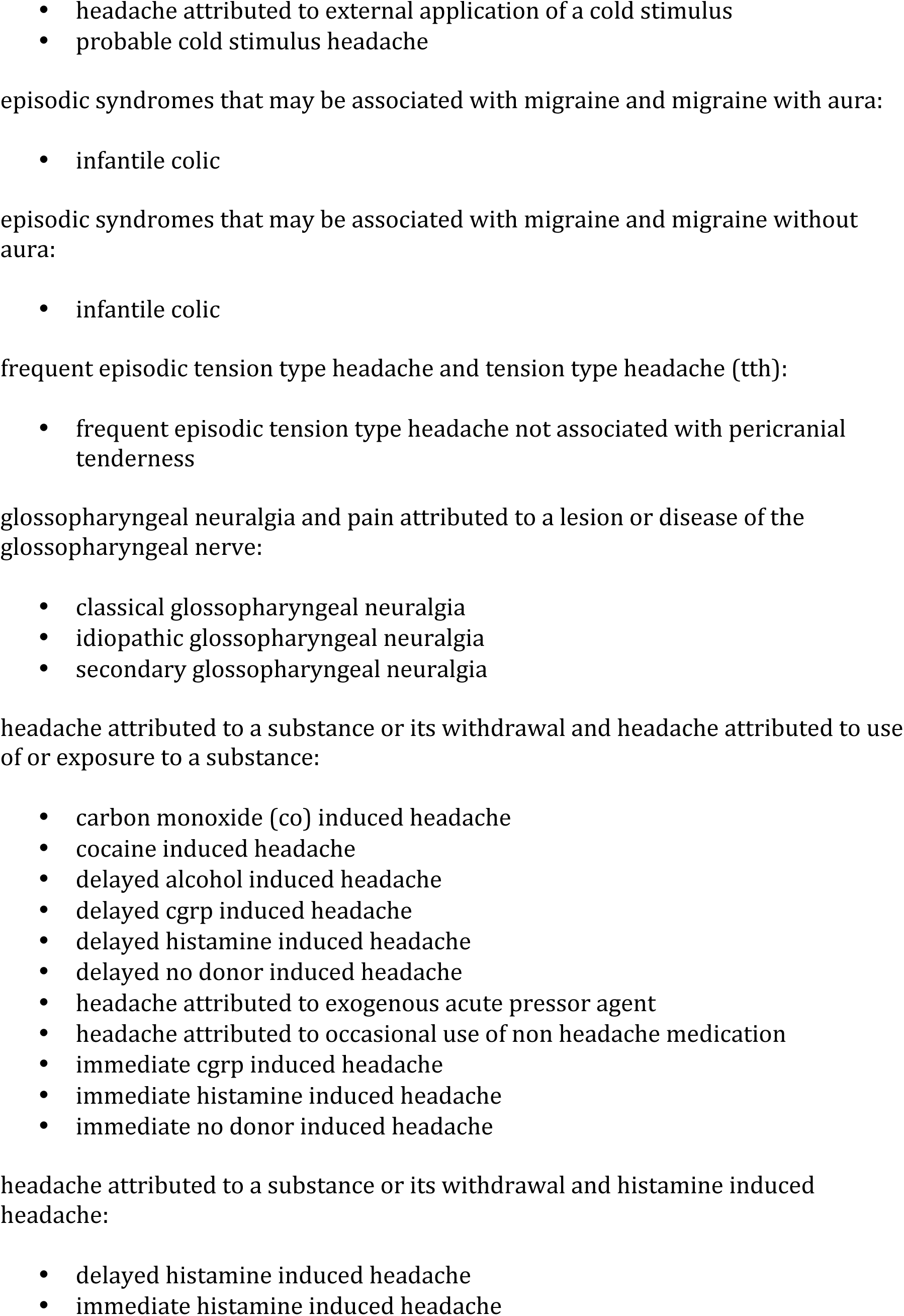
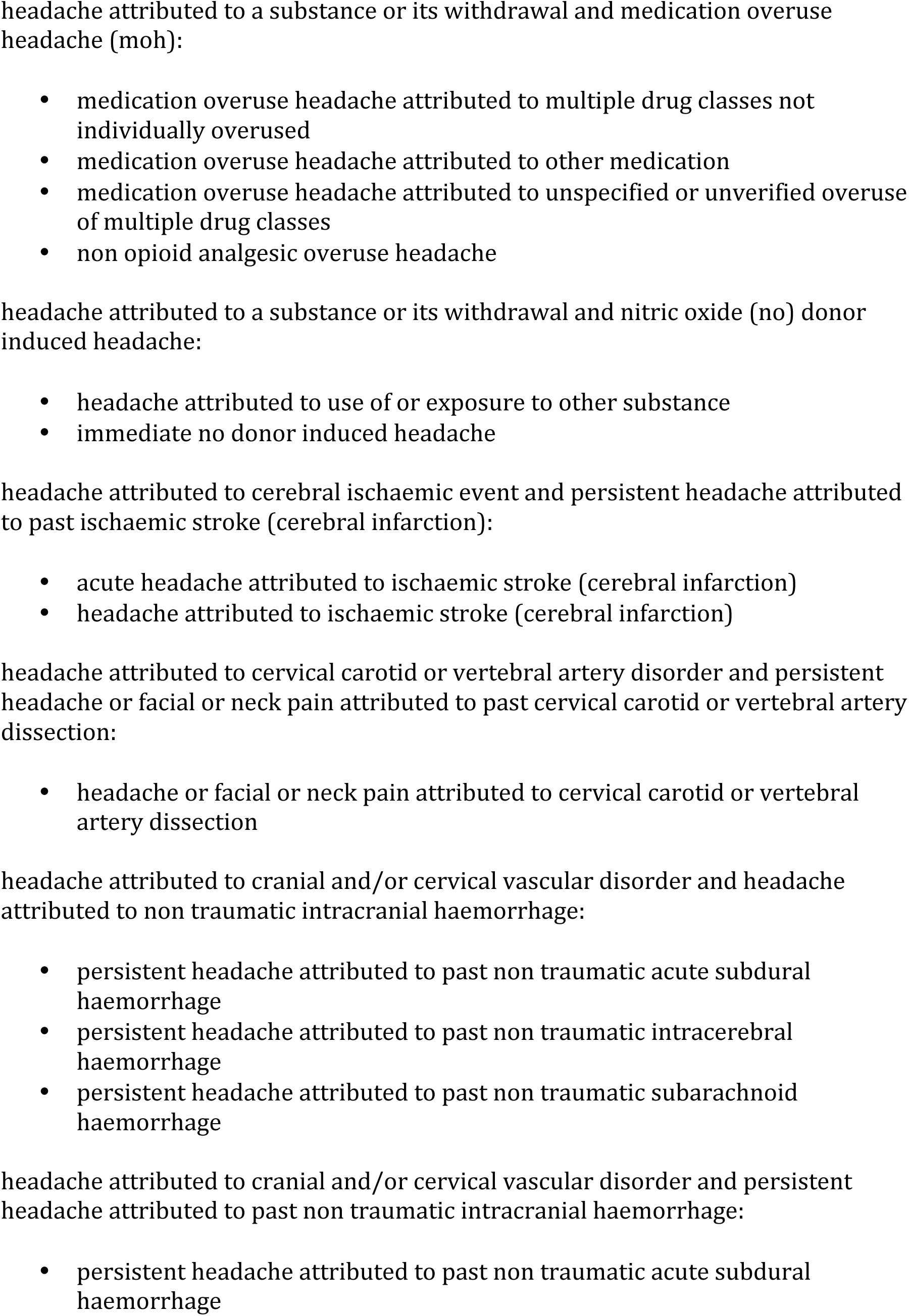
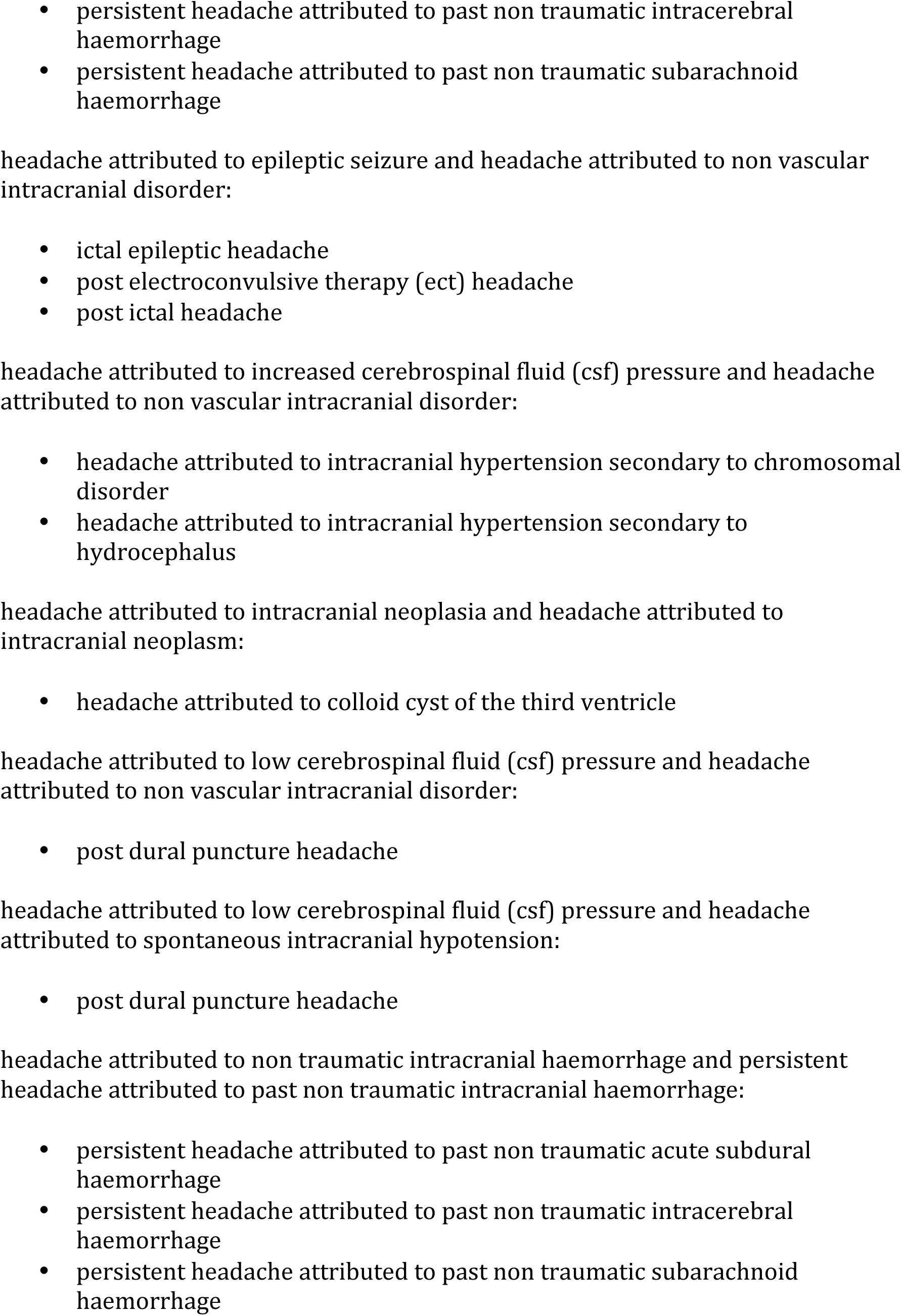
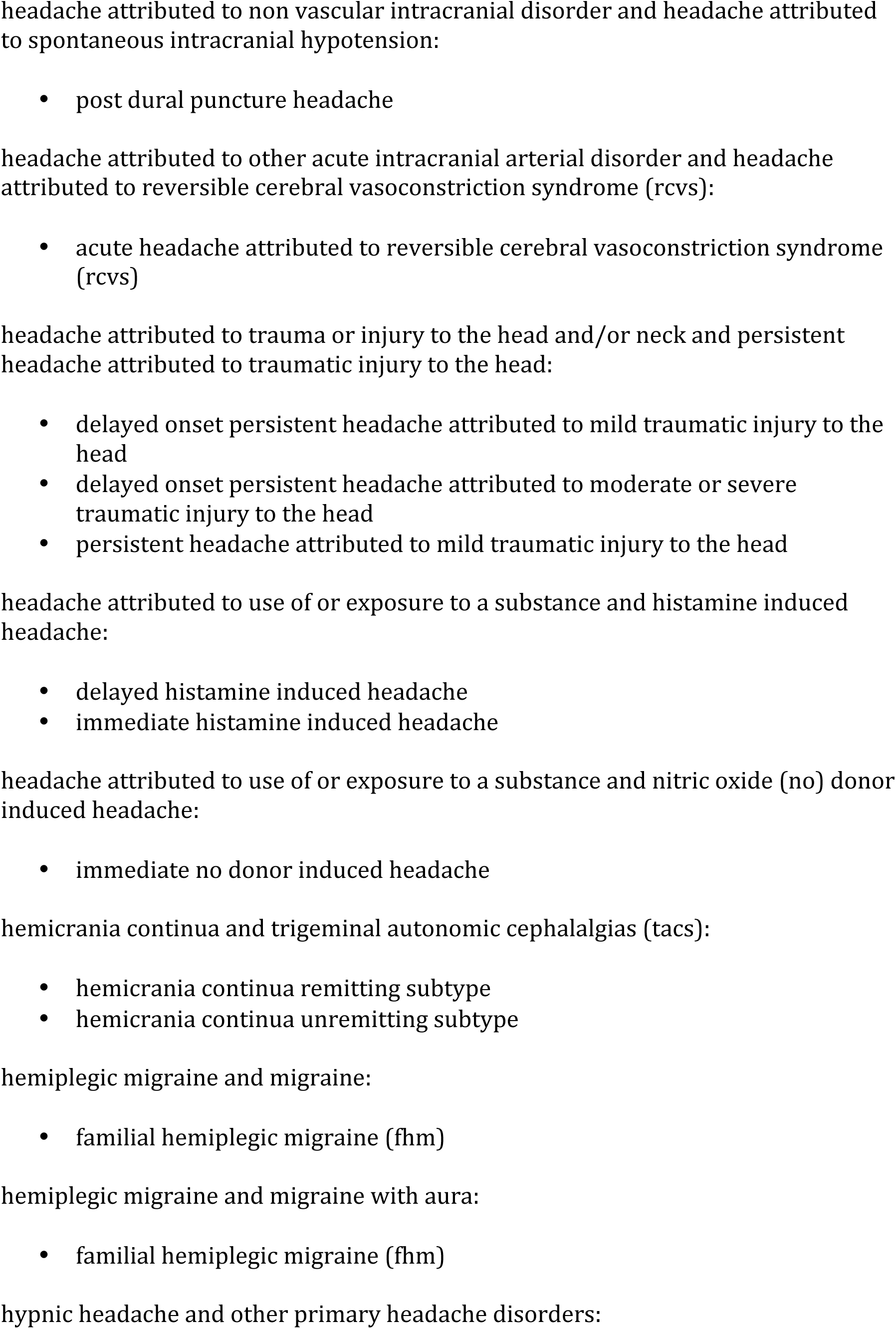
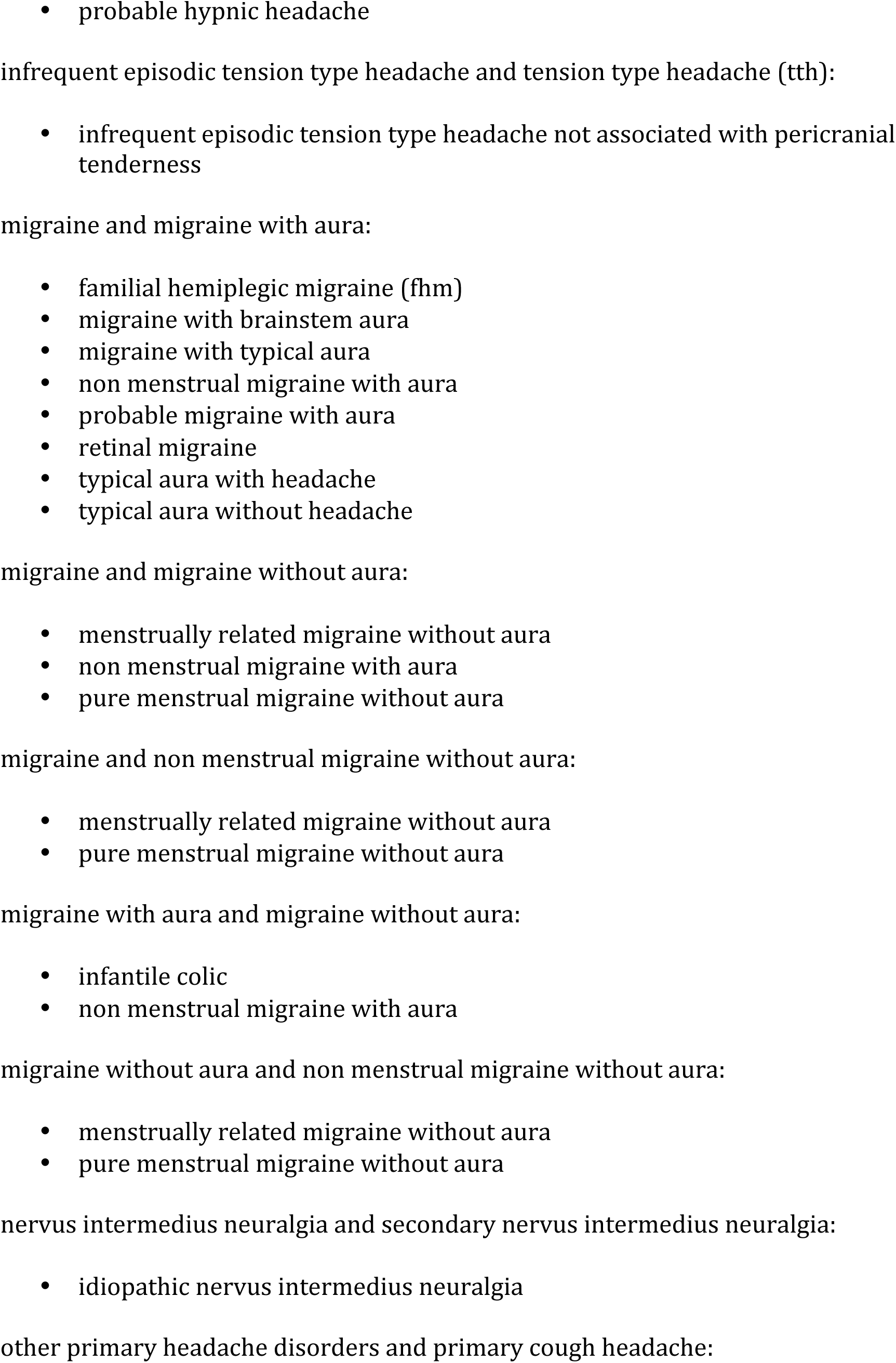
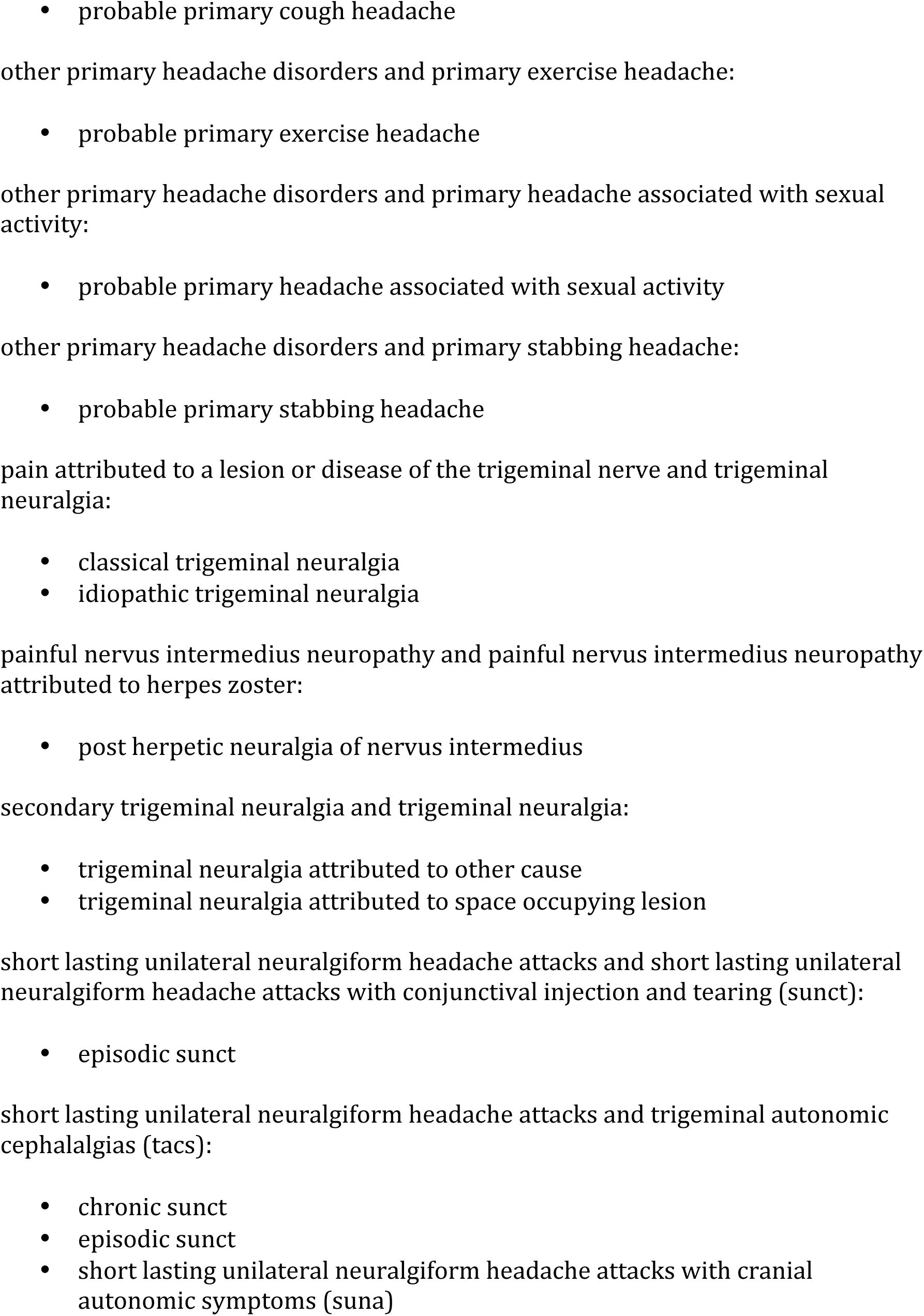
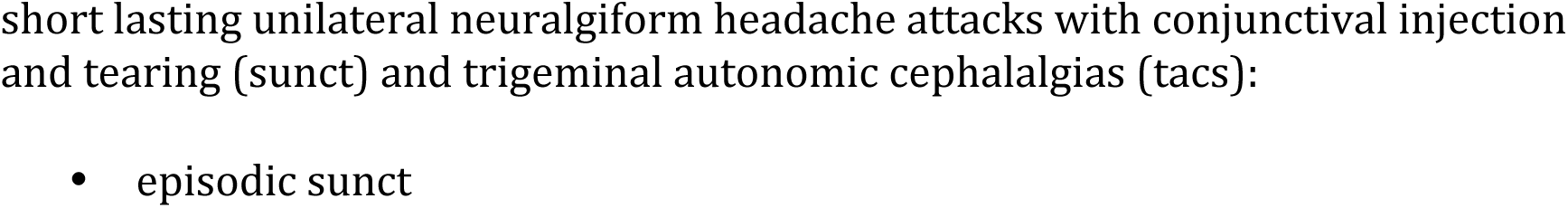
All possible “lower bound”

## Discussion

The goal of this project is two-fold: 1) to explore the methodological possibility of structuring subset relationship between differential diagnoses and 2) to apply this technique to headache disorders as presented in ICHD3. We believe such a construction for differential diagnoses carries the practical implications of allowing clinicians to widen or narrow differential diagnosis for real life scenarios in headaches; indeed, Table 2 of this project is intended as such a reference guide. As a methodological exploration, we envision the methods of this project to be applied to other differential diagnosis generators. The theoretical/mathematical implications of this construction are explored in the technical addendum.

(An important caveat: We must remember in dealing with these differential sets to not think of only the disease entity the differential set is named after, but rather its underlying set of differential diagnosis, which is what they are in reality.)

### Difference and Similarities with ICHD3 and its implications

To answer the question “Which sets of differential diagnoses are subsets of each other?” we consider the similarities and differences between ICHD3 classification and differential diagnosis hierarchy.

A number of ICHD3 diagnoses hierarchy do contain value for differential diagnosis generation. For example, when diagnosing a patient with “migraine with aura”, the ICHD3 allows practitioners to effectively classify and therefore generate a potential differential diagnosis for the specific types of migraine aura. Therefore, a number of super-sets are mirrors their ICHD3 counterparts:

Headache attributed to homeostasis (super-set 12)
Headache attributed to infection (super-set 15)
Headache attributed to non-vascular intracranial disorder (super-set 18)
Headache attributed to psychiatric disorder (super-set 19)
Headache or face pain attributed to disorder of the cranium, neck, eyes, ears, nose, sinuses, teeth, mouth, or other facial or cervical structure (super-set 24)
Painful lesions of the cranial nerves and other facial pain (super-set 38)
Trigeminal neuralgia (super-set 51)

Intuitively, the phenotypes within these sets reflect the underlying pathophysiology. For example, diseases from differet homeostasis may present phenotypically similar and therefore its diagnostic investigation (i.e. differential diagnoses) mirrors its classification.

Despite these similarities, the hierarchy generated by this project is in general “wider” yet “shallower” hierarchically than the ICHD3. For example, the maximum depth of ICHD3 is 5 levels (cf ICHD3 8.2.3.2.1 “acetylsalicyclic acid over-use headache”), where as the differential diagnosis hierarchy is only maximumly 4 levels deep. On the other hand, there are 3 parts with a total of 14 categories in ICHD3; our project requires a total of 93 differential diagnosis sets. In other words, differential diagnosis hierarchy splits disease classifications hierarchy. To understand the way in which this is accomplished, consider the following non-exhaustive list of differences between the hierarchy of ICHD3 and the hierarchy of differential diagnosis sets:

First, a number of stroke sub-classifications in ICHD3 become disparate super-sets in the differential hierarchy – for example, differential super-set 2, 17, 20, 39, and 40 are not subsumed under super-set 11, “headache attributed to cranial and/or cervical vascular disorder”. The likely explanation for this is that 2, 17, 20, 39, 40 are vascular disorders with symptoms that are phenotypically (although not necessarily pathophysiologically) different than the typical super-set 11 presentation. Consider RCVS (super-set 20) for example, the thunderclap onset of RCVS is distinct from what headaches from ischemic stroke classically presents. (Per ICHD3’s own admission, ischemic stroke headache presents in ischemic stroke in only one-third of the cases whereas the phenotype of RCVS as thunderclap made it into the criteria itself.) The immediate implication of this observation clinically is that clinicians should view vascular headaches as having 6 different phenotypes of presentations.

Similarly, traumatic headaches are described by super-set 1, 22, and 41. The likely etiology for this difference is that phenotypically acute versus persistence of headache post-traumatically represents two very distinct phenotypes in post-traumatic headaches. (We are not, of course, stipulating whether this is a correct/adequate classification of post-traumatic headaches, as is being questioned by a number of recent researches; we are simply stating that according to ICHD3, the acute versus chronic phenotype appears to be salient distinction between ICHD3 subtypes of post-traumatic headaches.)^9^

ICHD3 classification of “other primary headache disorders” (super-set 38) contains a number of headache conditions are so different that each becomes their own super-sets of differential diagnosis. This is evidenced by the fact that super-set 8, 28, 42, 43, 44, 45 ought to be included in super-set 38. Likely the reason is that although super-set 38 contains these diagnoses due to ICHD3 hierarchy, yet these diagnoses are so disparate that they simply contain their own sets of differentials.

High or low CSF pressure headaches, although pathophysiologically linked, present differently that their differentials form separate sets (super-sets 14, 16, 21). This ought not to be a surprise to any clinicians as elevated CSF pressure is associated with symptoms such as papilledema and vision changes while low CSF pressure is associated symptoms such as orthostatic headache.^5^ (Albeit apart from these textbook presentation, paradoxical cases exists.)^10^

Finally, we note that migraine is classified under various different super-sets: 26, 31, 32, 33, and 35. This highlights that phenotypically migraine without aura presents differently than migraine with aura (namely, the aura phase). Thus the aura phase induces very different clinical considerations in differential differential.

### Inversion of orders and violators of ICHD3 orders

Only a handful of differential diagnosis sets violates ICHD3 hierarchical order, as presented in Table 3. For the most part, these “violators” of ICHD3 hierarchy are simply minor restructuring of closely related ICHD3 diagnosis. Two of these “violators”, for example, are putting differential sets induced by “idiopathic” diagnosis under “secondary” diagnosis: for example, differential set induced by idiopathic nervus intermedius neuralgia is a subset of secondary nervus intermedius neuralgia. This simply reflects the reality of differential diagnosis generation: differential for considering secondary cause should be “bigger” than the differential of idiopathic causes for a disease, where presumably one considers the latter only when a number of known causes have been ruled out.

### 51 Super-set and the 42 Singletons

Our project offers an answer to the questions: “What are the minimum number of differential diagnosis sets that needs to be obtained in order to cover all of ICHD3 diagnosis?” We present the answer in Table 1 as 93 sets, differential diagnoses of which encompass all of ICHD3 diagnoses. An important clinical implication is that since the 93 lower-sets allows for a complete differential diagnosis of all of ICHD3 differentials, to efficiently diagnose patients clinicians may consider as first point of entry which of the 93 diagnoses a case is likely to represent.

There are 42 differential sets which we may call “singletons” – they are differential diagnoses sets that are neither subset nor super-set for other sets in the ICHD3. In other words, the diagnosis which induce the differential sets are unique. Some of these singleton sets appear to exist because their headache phenotype is so stereotypical so as to provide a very narrow differential. For example, MELAS and trochlear headaches are rather specific in that the former contains stereotypical image findings while the latter contains a stereotypical exam finding.^11, 12^ Others singletons appear to stand alone because the differential for them is broad in unique combinations. An example is NDPH, which provides a rather broad but unique differentials.^13^ “Probable migraine” and “probable tension type headache”, for example, likely offer similar broad differential; for the former, it is often made, only partly in jest, that “Everything is migraine… Unless It Isn’t.”^14^

### Strength

Methodologically, the ability to explore differential diagnosis as subset relationships allows for a well-structured classification of clinical reasoning in headache medicine. Such a structured view of differential diagnosis in headaches may allow clinicians to expand/narrow their differential structurally by tracing differential diagnosis hierarchy. As such, we hope that Table 2 would become for clinicians a useful resource. Since similarities or differences in differential diagnosis set is analogous to similarities or differences in disease presentation/phenotype, our classification can be viewed as a phenomenological classification of headaches (as opposed to the pathophysiological classification of ICHD3). Finally, since subset relationships satisfies what is mathematically termed a pre-order, when multiple clinical diagnoses are considered, sets can be “composed” additively, while still remaining theoretically coherent. (See theoretical addendum.)

Differential diagnosis generators can be variable in its quality. Using DiffNet as a source to study differential diagnosis sets has the benefits of being fully transparent in how differential diagnosis are generated. Specifically, there is a well defined way of enumerating differential for any given ICHD3 disorder under DiffNet. In addition, as DiffNet is simply a graphic representation of ICHD3, it is fully reproducible. As references between ICHD3 diagnoses are written by groups of international headache specialists, the source material for DiffNet is also of good authority. Finally, it has the benefit of being free to the public.

### Limitations and Future Direction

This project’s reliance on DiffNet and its methodology introduces limitations. Of course, cross-references in ICHD3 was not written for the purpose of differential diagnosis generation. As such, cross-references may not be clinical but pathophysiological, or at times maybe made to emphasize differences rather than similarities. This is an important limitation of using DiffNet for our project.

Furthermore, our prior editorial decision to include ICHD3 “alternative diagnosis” in DiffNet has downstream effect on this project: migraine or tension type headaches are presented with multiple definitions and, as such, comes with differential sets of differential diagnosis. A future direction should be to explore the effect of reconciling these differences.

Of course, methodologically, there is no reason why another differential diagnosis algorithm cannot be used to generate a differential diagnosis hierarchy using our methodology. Indeed, we encourage such an enterprise by readers to explore what would happen to differential diagnosis hierarchy if a neurology textbook, an alternative commercial project, or even an internal medicine textbook is used as input rather than DiffNet.

## Conclusion

We believe our projects carries implications for headache classification, for headache education, and finally as a differential diagnosis reference tool. For headache classification, our schema suggests that future headache classification schema may consider separations of a number of phenotypically different diseases. For headache education, we believe that education specifically on the 93 differential diagnosis supraset would provide a more comprehensive headache curriculum. Finally, we believe the classification schema in our project would serve as an effective differential diagnosis tool for clinicians.

## Clinical Implications

- Sets of differential diagnosis can be arranged in subset relationships for ICHD3 diagnosis. We offer this as a reference tool for the clinicians.
- The differential diagnoses from 93 ICHD3 diagnoses are sufficient to encompass all off ICHD3 diagnosis codes.

## Addendum 1 (The Technical Addendum)

The study of sets and their subsets relations belong to the mathematical discipline of order theory and lattice theory.^15^ The theoretical underpinning of this project is the application of these theory to the realm of differential diagnosis. In this technical addendum we will identify the mathematical and theoretical basis of our project.

First, let us introduce two closely related mathematical concepts, partial ordered sets (called posets for short) and preorder.^16, 17^

### Definition: Partial Preorder

Given P and elements a, b in P, then P is a partial preorder if:

1. a≤a for all a in P
2. If a≤b and b≤c then a≤c.

### Definition: Strict Preorder

1. (a < a) is false for all a in P
2. If a < b, and b < c, then a < c.

With the above two definitions, we can introduce partial ordered set, often called “poset” for short.

### Definition

Partial ordered sets is a set (P, ≤) such that P is a set and ≤ is a partial preorder on P.

Of note, any poset can be graphically represented using Hasse Diagram where set/subset relations are described by lines representing set/subset relationships in a downward fashion. We have avoided this presentation for clarity for the non-technical reader.

### A Potential Topological Implication

It is well established that posets is isomorphic to a topological space called the Alexandrof Space. Therefore, an implication of our project is that the subset relations between differential diagnoses may, with slight modification, be interpreted topologically.^18^ To our knowledge, this is only the second incidence of interpreting clinical entities topologically. (The first one was proposed by the author.)^19^ This correlation to topology can be exploited in future endeavors of translating theorems proved in Alexandrof Topology to the subset relation of differential diagnoses.

### Joint Semilattice

As a poset, our project turns differential diagnosis sets into a join-semilattice if we allow all 51 super-set as well as singletons to be subsumed under a common heading. (We can call this 1, as per lattice theory parlance.) Consider the following definition:

### Definition:^20^

Let (P, ≤) be a poset and x and y elements of P. Then meet of x and y, denoted x □ y, is the following, if it exists:

max{for all w in P: w ≤ x, w≤y}

Similarly, the join of x and y, denoted x □ y, is the following, if it exists:

min{for all z in P: x ≤ z, y≤z}

In other words, meet is the maximum lower bound and join is the minimum upper bound between x and y. We can then define semilattice as the following: If every pair in x, y in P has a join then it forms a join semilattice. Similarly, if every pair x, y in P has a meet then it forms a meet semilattice.^21, 22^

We can prove computationally that our 51 poset is indeed such a construction. (We include this as Supplementary Material 1.) Since the join of any two singleton is 1 and the join of any singleton with any elements of the 51 poset semilattice is also 1, we can then confirm that our construction forms a join semilattice.

**“Addition” of Differential Diagnosis as Algebraic implication**

Any join semi-lattice can be viewed as a idemopotent and commutative monoid under “join” operation, therefore our construction is a monoid.^23^ We can view the “join” operation as therefore a sort of “addition” for differential diagnoses. For example, given differential diagnosis a and b, we can always find a differential diagnosis set that includes both as its subset. For example, the “addition” (under “join” operation) of differential sets “visual snow” (31v) and “retinal migraine”(31r) would be the differential diagnosis set of “migraine”(31).

More interestingly is investigating whether “meet” exists in our poset; if addition of differential diagnosis through “join” is considered “expansion” of differential, then addition of differential through “meet” would be consider the narrowing of differential. We screened all possible non-trivial lower bound for meet as part of our project. (Trivial here means that if a = meet(a, b) then it is trivial.) Result is presented in Table 4. Unfortunately, the set of meet is not surprising, as the majority of meet are actually obvious connections between two diseases.

### Categorification and implication for diagnostic rule out

Our construction of subset relations between differential diagnosis can be easily turned into a strict poset relationship if we uses ≤ rather than <. If we allow for such a simple modification of our project, then our construction for differential diagnosis conveniently forms a mathematical category. This is due to the fact that any poset forms a thin category. We introduce both of these definitions below:

### Definition:^24, 25^

A category A consists of:

1. A collection of objects, denoted obj(A)
2. For each a, b in obj(A), a collection of maps, also called arrows or morphism, from a to b, denoted hom_A_(a, b). Often written as hom(a, b). In other words, per MacLane:

hom_A_(a, b) = {f | f is an arrow f: a -> b in A}
3. For each a, b, c in hom(A), a function hom(b, c) × hom(a, b) -> hom(a, c) called composition.
4. For each element a of obj(A), an element 1_A_ in hom(a, a), called identity on a.

The above data must satisfy the following:

1. Associativity: for each f in hom(a, b) and g in hom(b, c) and h in hom(c, d) then (h . g) . f = h . (g . f)
2. Identity: for each f in hom(A, b), f . 1_A_ = 1_B_ . f.

A thin category is a category with at most one morphism. It is also called a posetal category.

### Definition:^22^

Let C, B be two categories. A functor is defined by two functions: and object function T and an arrow function, also written as T, satisfying the following:

1. Object function T assigns an each object c in C to an object Tc in B.
2. Arrow function assigns each arrows f in hom(c, c’) in C to an arrow Tf in hom(Tc, Tc’) of B.
3. T(1_c_) = 1_TC_
4. T(g . f) = Tg . Tf, whenever composition is defined in C for g and f.

Establishing functorial relationships between different categories allows us to describe diagnostic rule outs. For example, consider the case where “headache secondary to TIA” is ruled out by a diagnostic intervention. We can construct a new poset from our existing one by simply taking out “headache secondary to TIA” from all of its elements. (Represented by the notation “\”.) One can show that the new poset constructed through this rule out retains the exact same set/subset relationship.

Diagnostic rule out of “headache secondary to TIA” from our original poset is therefore a functor in the following way 1) the functor maps differential set in the original poset to a differential set without the “headache secondary to TIA” and 2) the functor maps the function function ≤ to ≤ in the new poset. That is, F(a ≤ b) = Fa ≤ Fb. Notice that F(f . g) = F (x < z) = (x \ b) ≤ (z \ b) for any f: x-> y and g: y -> z, where b = “headache secondary to TIA”. Also F(f) . F(g) = (x \ b ≤ y \ b) ≤ ( y \ b ≤ z \ b) = (x \ b) ≤ (z \ b). Therefore the functor relationship is satisfied.

### Theoretical Conclusion and Future directions

Our project provides a potential theoretical foundation for the exploration of orders and hierarchy in clinical science apart from ordering based on pathophsyiology. In other words, while ICHD3 classification classifies headache disorders pathophsyiologically, our schema establishes a foundation for classification of differential diagnosis based on established mathematical framework.

## Conflict of Interest Statement

Pengfei Zhang: He has received honorarium from Alder Biopharmaceuticals, Board Vitals, and Fieve Clinical Research. He collaborates with Headache Science Incorporated without receiving financial support. He has ownership interest in Cymbeline LLC.

## Financial Support

This research received no specific grant from any funding agency in the public, commercial, or not-for-profit sectors.

## Trial Registration

Not applicable.

## Abbreviations

## Supporting information

Supplement Proof

## Data Availability

All data produced in the present study are available upon reasonable request to the authors

## Acknowledgements

None

## References

1. Scordo KA. Differential diagnosis: correctly putting the pieces of the puzzle together.AACN Adv Crit Care.;25:230–6.

2. Nardone DA. Differential diagnosis and heuristics.JAMA.;254:2890–1.

3. Zhang P. DiffNet. http://techfinder.rutgers.edu/tech/DIFFNET. Accessed October 26, 2021.

4. Zhang P, Berk T. “IHC 2019 Abstracts: Network Analysis of the International Classification of Headache Disorders, 3rd Edition” Cephalalgia 39:Supplement: 1-337.

5. The International Classification of Headache Disorders, 3rd Edition. Cephalalgia 2018;38:1-211.

6. Weisstein, Eric W. “Set”. mathworld.wolfram.com. Retrieved 2020-10-27

7. Weisstein, Eric W. “Subset”. mathworld.wolfram.com. Retrieved 2020-10-27

8. Weisstein, Eric W. “Superset”. mathworld.wolfram.com. Retrieved 2020-10-27

9. Ashina H, Eigenbrodt AK, Seifert T, Sinclair AJ, Scher AI, Schytz HW, Lee MJ, De Icco R, Finkel AG, Ashina M. Post-traumatic headache attributed to traumatic brain injury: classification, clinical characteristics, and treatment.Lancet Neurol.2021;20:460–469.

10. Cheng SJ, Hakkinen I, Zhang P, Roychowdhury S. Paradoxical headache in a case of chronic spontaneous intracranial hypotension and multiple perineural cysts.Headache.2021;61:1291–1294.

11. Malhotra K, Liebeskind DS. Imaging of MELAS.Curr Pain Headache Rep.2016;20:54.

12. Tran TM, McClelland CM, Lee MS. Diagnosis and Management of Trochleodynia, Trochleitis, and Trochlear Headache.Front Neurol.2019;10:361.

13. Evans RW. New daily persistent headache.Curr Pain Headache Rep.2003;7:303–7.

14. Purdy A. 19th Annual HCNE Boston Fall Headache Symposium. Nov 2, 2019.

15. Gratzer G. Two Problems That Shaped a Century of Lattice Theory. Notice of the AMS. 54;6:696–707.

16. Roitman J. Introduction to Modern Set Theory. 3rd ed. Orthogonal Publishing; 2013.

17. Fong B, Spivak D. An Invitation to Applied Category Theory: Seven Sketches in Compositionality. Cambridge University Press: 2019.

18. Arenas FG. Alexandroff Spaces. Acta Math. Univ. Comenianae. 1999: 17–25.

19. Zhang, P “Categorical Medicine: A Mathematical Embedding of Clinical Medicine with Applications to Headache Medicine.” OSF Preprints. September 13. doi:10.31219/osf.io/xw6t5.

20. Jongsma, C. Discrete Mathematics: Chapter 7, Posets, Lattices, & Boolean Algebra. 2016: Retrieved from https://digitalcollections.dordt.edu/faculty_work/427 Accessed 10/29/2021

21. Ditor SZ. Cardinality Question Concerning Semilattices of Finite Breadth. Discrete Mathematics 1984(48):47–59

22. Gratzer G. General Lattice Theory. Birkhauser Verlag; 2003.

23. . Friedrich Wehrung. Refinement monoids, equidecomposability types, and Boolean inverse semigroups. Springer Verlag, 2188, 2017, Lecture Notes in Mathematics, 978-3-319-61598-1. ff10.1007/978-3-319-61599-8ff. ffhal-01197354v3f

24. Leinster T. Basic Category Theory. Cambridge University Press: 2014.

25. MacLane S. Category Theory for the Working Mathematician. Springer: 1971.

